# Reducing Exposures to Airborne Particles Through Improved Filtration: A High-Level Modeling Analysis

**DOI:** 10.1101/2020.05.14.20101311

**Authors:** Michael B Dillon, Richard G Sextro

## Abstract

Indoor airborne particulates are well-known health hazards and filtration is one common method of reducing exposures. Based on our previously developed Regional Shelter Analysis methodology and parameters that characterize the existing US building stock, we perform a high-level assessment of the potential benefits of upgrading existing filters in furnace and in heating, ventilation, and air conditioning systems using off-the-shelf filters. We use three metrics to assess the improvement: Building Transmission Factor (a measure of protection against outdoor airborne particles), Indoor Normalized Time and Space Integrated Air Concentration (a measure of indoor exposure to indoor-origin airborne particles), and Building Exit Fraction (fraction of indoor airborne particles that are released to the outdoor atmosphere). We also discuss the potential reduction in regional exposures due to particles exiting the building and exposing downwind building occupants.

Our modeling indicates that while buildings provide their occupants passive protection against airborne particulate hazards, including but not limited to PM_2.5_, PM_10_, and wildfire smoke; improving particle filtration efficiency may further improve this protection. The degree of improvement varies with particle size and building type. Of the building types studied, apartments are predicted to benefit most, with greater than a factor of 2 improvement (≥50% reduction in exposures) for 1 µm particle exposures when using MERV 7 to 12 rated filters. Non-residential buildings were notably less responsive to improved filtration but had the highest Building Exit Fractions with 30% to 40% of indoor airborne particles released to the outdoor atmosphere (apartment buildings only released 6% to 9%). Improvements predicted for single family homes were intermediate between apartments and non-residential buildings. Improvements in the Regional Exposure metric are larger, ranging from a factor of 2.5x to 10x for residences (when using MERV 7 to 12 rated filters) and up to 25x for large apartments with MERV 14 or 15 rated filters. The results of our modeling analysis are broadly consistent with the limited experimental data and modeling results available in the literature.

## 2. Introduction

Indoor exposure to airborne particles can arise from many outdoor and indoor sources and sufficient exposures are known to result in acute and chronic lung disease as well as heart disease [1]–[5]. Buildings can passively provide – in some cases significant – protection to their occupants from a broad range of particle inhalation hazards – including but not limited to PM_2.5_, PM_10_, and wildfire smoke – as discussed in detail in [6], [7]. Using the Regional Shelter Analysis (RSA) method described in [6], we previously estimated the passive protection provided by the existing US building stock for airborne particulate hazards [7]. The present work – utilizing the same RSA approach – focuses on the degree to which straightforward changes in building operation could yield an additional measure of protection. Specifically, this report considers the building protection benefits of improving current particle filtration efficiency by (a) using off-the-shelf furnace and heating, ventilation, and air conditioning (HVAC) system filters and (b) increased furnace fan operation for residential buildings.

Our modeling uses the Building Transmission Factor metric (BTF) which is the ratio of indoor to outdoor exposures, where exposures are time-integrated airborne particle concentrations. The BTF, sometimes called the Building Exposure Ratio, is the inverse of the Building Protection Factor metric used in our previous RSA work [6], [7]. In addition to the BTF, two related, but distinct, building metrics are also of interest and are analyzed here: (a) the Indoor Normalized Time and Space Integrated Air Concentration (TSIAC) which is the degree to which indoor individuals are exposed to indoor airborne particles (the indoor time and space integrated air concentration normalized to the release of a unit amount of airborne material, such as a single particle or a µg of material) and (b) the Building Exit Faction (BEF) which is the fraction of indoor airborne particulates that exit the building and enter the outdoor atmosphere. The potential improvement in (i.e., reduction in) regional exposures that arises due to the reduction in both the number of particles exiting the building and entering downwind buildings is also discussed.

## 3. Building Models and Key Metrics

Outdoor airborne particles can enter a building through mechanical ventilation (e.g., HVAC systems), natural ventilation (e.g., open windows), and/or infiltration (e.g., through exterior wall cracks). Once indoors, airborne particles can be removed from the indoor air through (a) air leaving the building by mechanical or natural ventilation and exfiltration; (b) active filtration by recirculation through furnace or HVAC systems (if present); (c) deposition on indoor surfaces (particle resuspension is not accounted for in this study); and (d) other processes, including but not limited to, chemical reactions and portable (stand-alone) indoor air filtration systems (neither of which are considered here). For this study, we have assumed building doors and windows are closed.

Modeling indoor contaminant concentrations requires choosing among a variety of mathematical models with increasing complexity, ranging from simple single compartment models to multizone models to highly detailed computational fluid dynamics models. While increasingly detailed and complex models may reduce modeling conservatism and uncertainty, the number and required fidelity of the input parameters also increases (see [8] for a general discussion). Since our goal is to provide a broad, high level assessment across a range of building categories and, as a practical matter, detailed parameters are not generally available for many buildings of interest; we make two key modeling assumptions (these assumptions are consistent with prior studies [9]–[12]). First, indoor air volumes, such as the breathing space of a building, can be represented as a single compartment, which can be used to describe the time evolution of indoor contaminant concentrations. Second, airborne particle concentrations within that single compartment are spatially uniform.

These assumptions are codified in the single box model (**Equation 1**) which can be used to describe the time evolution of indoor airborne particle concentrations. This study includes the additional, commonly used assumption that the transport and loss terms, i.e., the λ parameters, are independent of both time and air concentration on the timescales of interest.

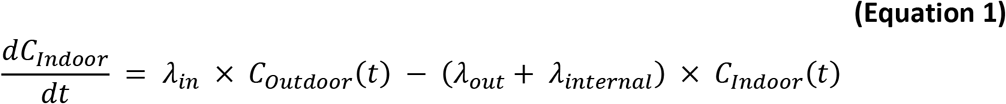

where

*λ_in_* = the rate at which outdoor airborne particles enters the building – typically via infiltration or ventilation. Includes losses that occur during transport from outdoor to indoor (h^−1^)
*λ_internal_* = the rate at which indoor airborne particles are lost within the building – typically by deposition to surfaces or by filtration (h^−1^)
λ_out_ = the rate at which indoor airborne particles exit the building – typically via exfiltration or ventilation (h^−1^)
*C_Indoor_*(t) = the indoor particle air concentration at time t (g m^−3^)
*C_Outdoor_*(t) = the outdoor particle air concentration at time t (g m^−3^)
*t* = time (h)

We adapt **Equation 1** to develop two sets of generalized equations for each of the three building metrics (Building Transmission Factor, Indoor Normalized TSIAC, and Building Exit Fraction). Each equation set is defined on the basis of one of two common combinations of (a) the two building airflow mechanisms described above (filtered recirculation; HVAC systems) and (b) the relevant indoor loss mechanisms. In addition to any filtration that might be present, and already incorporated into the airflow terms, we consider two additional indoor loss mechanisms: (a) deposition of airborne particles to indoor surfaces, λ_*dep*_, and (b) a generic first-order airborne loss mechanism that can be used to account for other loss mechanisms not explicitly considered in this analysis, λ_*decay*_. Several parameters are particle-size-dependent. For readability, these dependencies are not shown in the equations themselves, but are denoted by (particle size) in the list of variable definitions.

The first set of airflow equations (denoted by **R**) corresponds to buildings with filtered recirculation, which in the US are typically residences. Outdoor airborne material enters the building only through the infiltration pathway. The forced air furnace recirculation air filter, if present and active (fan on), removes a fraction of indoor airborne particles.

The second set of airflow equations (denoted by **H**) corresponds to buildings with an active HVAC system, which in the US are typically non-residential buildings. Outdoor airborne material enters the building through either infiltration or the HVAC system outdoor air intake. The HVAC system air filter removes airborne particles from both the entering and recirculating air. This equation implicitly assumes that the HVAC system fan duty cycle is 100% (the system is always moving building air, although not necessarily heating or cooling it).

### 3.1. Reducing Indoor Exposures to Outdoor Airborne Particles (Building Transmission Factor)

The building transmission factor equations were previously derived in [6], [7], where the Building Protection Factor metric was used. **Equation 2** provides the general form. This term is the ratio of the total indoor exposure to total outdoor exposure. For this application, exposure is defined as the time-integrated airborne particle concentration, hence the Building Transmission Factor is not simply the ratio of indoor to outdoor concentrations. [6], [7] provide more details, discussion, and alternative equations for other exposure metrics.

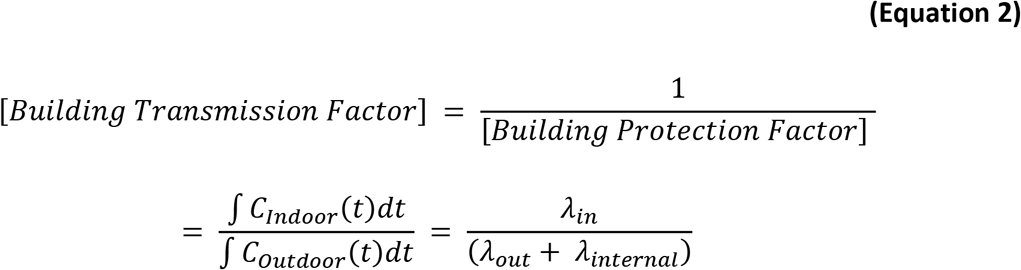

For buildings with filtered recirculation, **Equation 3R** specifies the Building Transmission Factor.

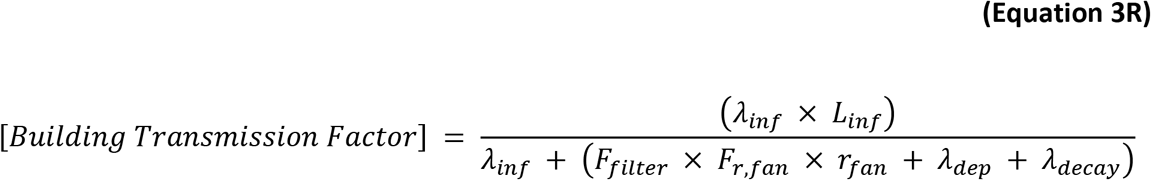

For buildings with an active HVAC system, **Equation 3H** specifies the Building Transmission Factor.

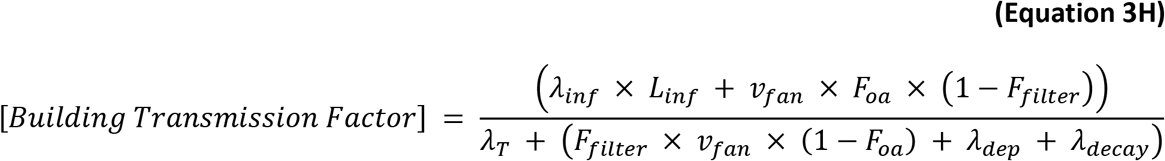

where

λ_dep_(particle size) = the particle-size-dependent indoor deposition loss rate (h^−1^)
λ_decay_ = the “generic” first-order airborne decay (loss) rate. This term is used to estimate the impact of loss mechanisms that are NOT otherwise specified. (h^−1^)
λ_inf_ = the air infiltration rate at which air enters a building (h^−1^)
λ_T_ = the total building ventilation rate [= sum of the infiltration and mechanical ventilation rates] (h^−1^).
L_inf_(particle size) = the particle-size-dependent efficiency by which particles can penetrate the building shell (dimensionless)
F_filter_(particle size) = the particle-size-dependent filtration efficiency (dimensionless)
F_oa_ = the fraction of outdoor air passing through the HVAC supply fan (dimensionless)
F_r,fan_ = the fraction of time the forced air furnace recirculation fan is on, i.e., the fan’s duty cycle (dimensionless)
r_fan_ = the rate at which a building volume of air recirculates through the furnace systems when the fan is on (h^−1^)
v_fan_ = the rate at which a ventilation or HVAC supply fan delivers a building volume of air when the fan is on and combines the outside and recirculation air rates (h^−1^)
[Building Protection Factor] = ratio of the outdoor to indoor exposure. Similar to sunscreen and personal protective respirator rating systems, higher protection factor values indicate lower exposures and thus increased protection. (protection factor)
[Building Transmission Factor] = ratio of the indoor to outdoor exposure. ((protection factor)^−1^)

### 3.2. Reducing Indoor Exposures to Indoor Airborne Particles (Indoor Normalized TSIAC)

The following equations define the indoor normalized time and space integrated airborne particle concentration (Indoor Normalized TSIAC). While this metric can be applied more generally, it is used here to evaluate the exposure to particles released indoors. The equations are developed with the additional assumptions that outdoor airborne particle concentrations are zero and the indoor emission of a single particle occurs at single time (t = 0). With these assumptions, **Equation 1** simplifies to **Equation 4**. When a unit amount of material is released indoors at t = 0, **Equation 4** reduces to **Equation 5**.

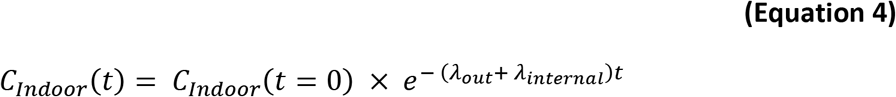

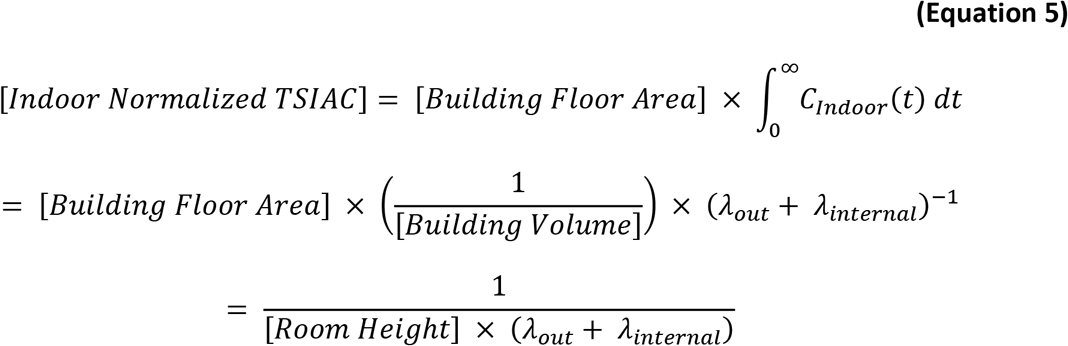

For buildings with filtered recirculation, **Equation 6R** specifies the Indoor Normalized TSIAC.

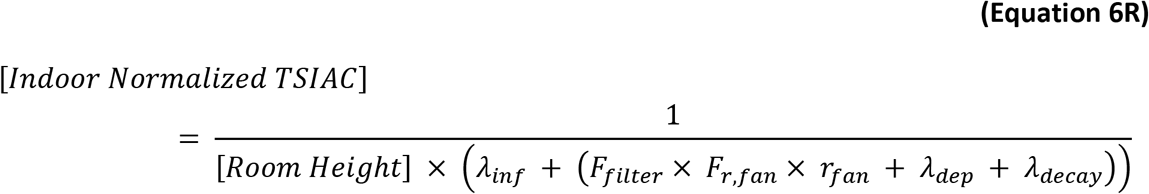

For buildings with an active HVAC system, **Equation 6H** specifies the Indoor Normalized TSIAC.

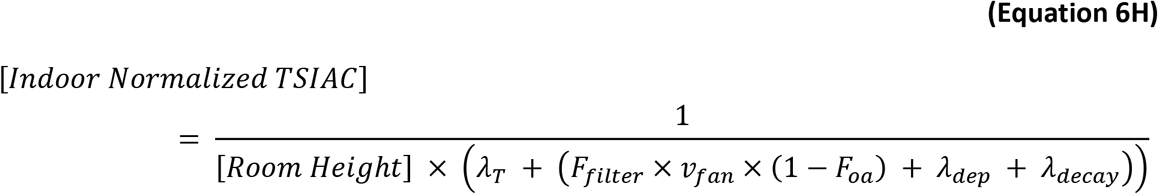

where

[Building Floor Area] = floor area of the building (m^2^)
[Indoor Normalized TSIAC] = indoor time and space integrated air concentration assuming a unit amount of material is released (s m^−1^)
[Room Height] = height of building occupied space (m)

### 3.3. Fraction of Indoor Airborne Particles Released to the Outdoors (Exit Fraction)

**Equation 7** shows the fraction of material released indoors that exits the building and enters the outdoor atmosphere. This equation is based on (a) the appropriate (normalized) indoor concentration (**Equation 5**), (b) the rate at which indoor air (and airborne material) leaves the building, and (c) particle losses to the building envelope that occur while airborne particles suspended in the indoor air are exiting the building.

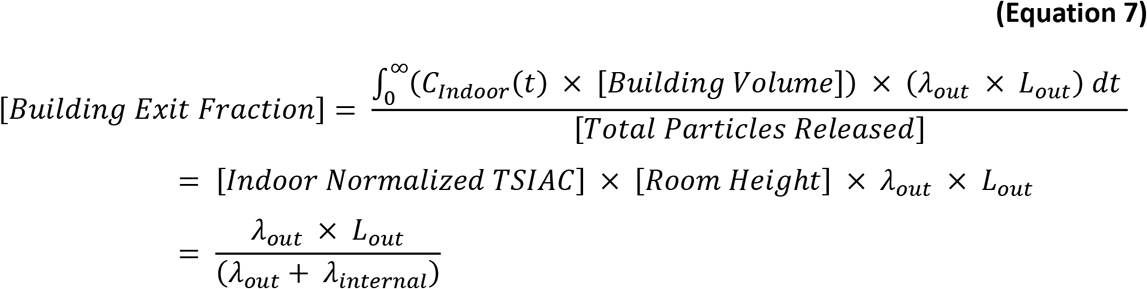

For buildings with filtered recirculation, outdoor air enters the building only through the infiltration pathway and so λ_out_ = λ_inf_ and **Equation 8R** specifies the building exit fraction. We assume here that most of the indoor air that exits the building does so via the exfiltration pathway (as opposed to other exit pathways such as bathroom or kitchen exhaust fans, and dryer vents) and therefore L_out_ = L_inf_.

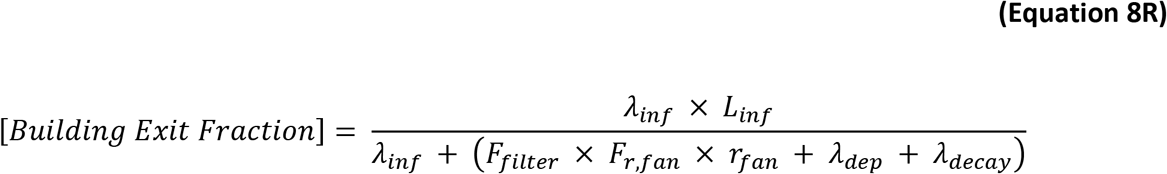

For buildings with an active HVAC system, indoor airborne material exits the building through either infiltration or through mechanical means, e.g., the HVAC system exhaust; **Equation 8H** specifies the building exit fraction. Again we note that the entrance and exit airflow paths may differ, but as a practical matter assume (a) that they are the same (λ_out_ = λ_inf_; L_out_ = L_inf_) and (b) no particles are lost while the indoor air is exiting through the exhaust system.

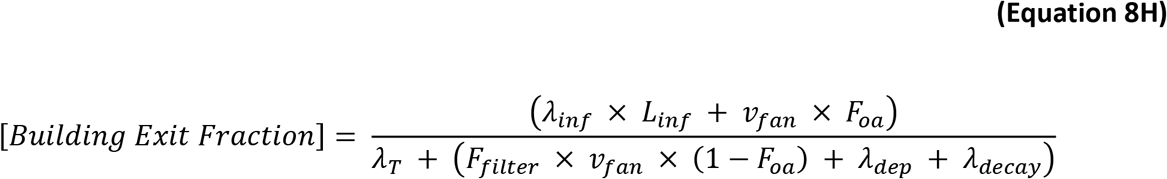

where

[Building Exit Fraction] = fraction of material released within a building that exits the building and enters the outdoor atmosphere. (no units)
L_out_(particle size) = the particle-size-dependent efficiency by which particles in indoor air exit the building through cracks and other penetrations in the building shell (dimensionless)

### 3.4. Regional Exposures

**Equation 9** describes the fraction of indoor airborne particles that exit a source building, are transported downwind, and enter another building’s indoor air space. This metric incorporates both the source Building Exit Fraction (the fraction of indoor airborne particles that exit the source building) and the downwind Building Transmission Factor (the fraction of outdoor airborne particles that enter the second building’s air space). The *[Outdoor Normalized TSIAC]* term in **Equation 9**, which requires analysis beyond that presented here, assesses the degree to which particles, once emitted from the source building, are transported and diluted in the outdoor atmosphere. **Equation 10** defines the Improvement in Downwind Indoor Exposure metric as a ratio to the baseline scenario and so the *[Outdoor Normalized TSIAC]* terms drops out. This metric can be used to assess the degree to which improved filtration scenarios decrease downwind indoor exposures relative to the baseline scenario.

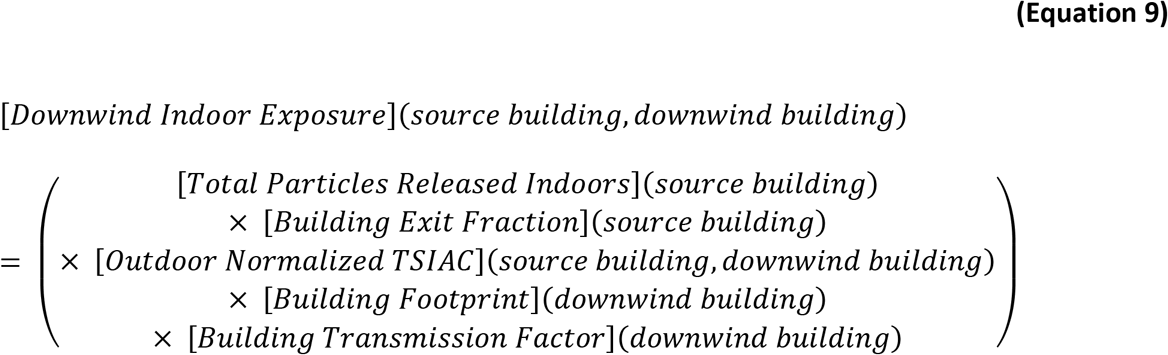

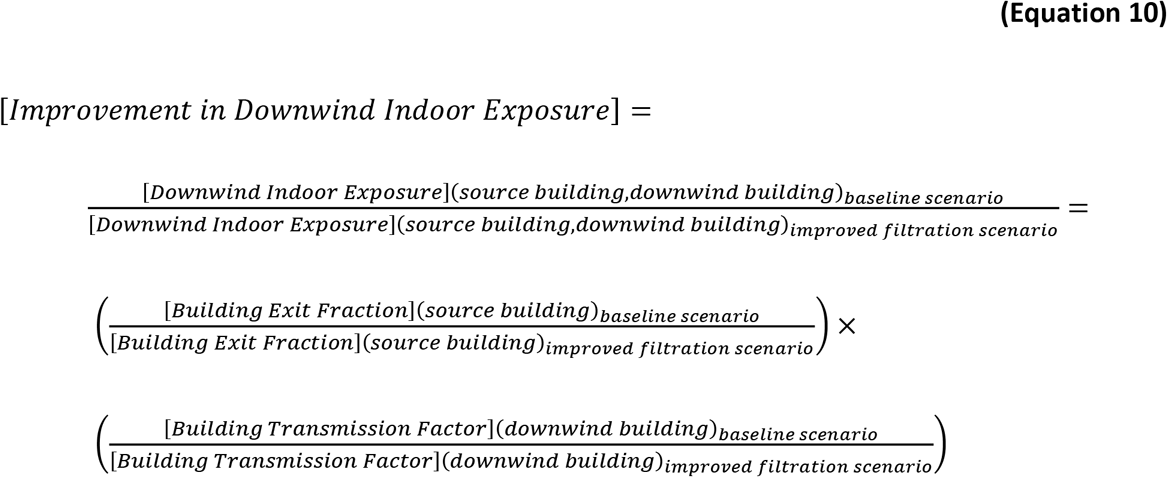

where

[Building Footprint](building) = the projected area of the building onto the Earth’s surface. (m^2^)
[Downwind Indoor Exposure](source building, downwind building) = the number of airborne particles in the breathing volume (respiratory second volume) of an individual in the downwind building that were emitted within the source building. (particles s m^−3^)
[Improvement in Downwind Indoor Exposure] = ratio of indoor downwind exposure for the baseline scenario to the improved filtration scenario. (no units)
[Outoor Normalized TSIAC](source building, downwind building) = particle air concentration outside the downwind building integrated over the passage of the airborne particulate plume and the building footprint assuming a single particle was released to the outdoor atmosphere from the source building. (s m^−3^)
[Total Particles Released Indoors](source building) = total number of airborne particles released within the source building. (particles)

## 4. Building Parameter Values

Our modeling is based on the building use type categories and parameters values described in our earlier report [7] and briefly summarized here. The tables that follow are adapted from that report, where additional discussion and descriptive details may be found. We provide building air flow and particle parameters for most of the US Department of Homeland Security (DHS) HAZUS convention of 33 building use types (termed building occupancy types in the HAZUS documentation), see **Table 1**; and for individuals outdoors [13]. These parameters are used when calculating the Building Transmission Factor, Indoor Normalized TSIAC, and Building Exit Fraction values from the **R** and **H** airflow equations.

To assess the benefits from increasing building filter efficiencies, this report compares the effects on indoor airborne particulate concentrations arising from the existing US building stock filter efficiencies – as a baseline scenario – to the concentrations yielded from using higher efficiency filtration scenarios that could be implemented. The only differences in parameters between the baseline scenario and the 3 improved filtration scenarios analyzed are (a) different furnace duty cycles (*F_r, fan_*) in **Table 3** below and (b) the choice of filtration categories in **Table 9** below. Room height, used in the calculation of the Indoor Normalized TSIAC metric, is assumed to be 3 m for all buildings.

### 4.1. Building Model Assignment

The **Table 2** column “Building airflow type” assigns the appropriate airflow equation type (**R** = filtered recirculation; **H** = HVAC system) to most US FEMA HAZUS building use type categories specified in **Table 1**, and also specifies the appropriate airflow parameter values associated with building infiltration, ventilation, and filtration. We note that the prior studies of building performance and/or indoor pollutant behavior (discussed below) were not performed to explicitly conform to HAZUS building use type categories. For some HAZUS building use type categories, e.g., RES1, the mapping between the prior work and the HAZUS category is straightforward. The mapping for other HAZUS categories is less clear and for some categories there are limited published data available. For these cases, we made estimates based on best engineering judgment and adapted the available input data to conform to the HAZUS building types (as discussed below and in [7]). Single box modeling may not adequately represent the indoor airborne particle concentrations for some parking garages (COM10) and so we exclude this building use type from our analysis.

**Table 1.** Mapping between HAZUS provided occupancy class and specific building use types. This information is adapted from Table 13.2 presented in [13].

| Occupancy class | Building use type | Building use type description |
| --- | --- | --- |
| **Residential** | RES1 | Single family dwelling |
|  | RES2 | Manufactured (Mobile) home |
|  | RES3A | Multi-family dwelling: duplex |
|  | RES3B | Multi-family dwelling: 3–4 units |
|  | RES3C | Multi-family dwelling: 5–9 units |
|  | RES3D | Multi-family dwelling: 10–19 units |
|  | RES3E | Multi-family dwelling: 20–49 units |
|  | RES3F | Multi-family dwelling: 50+ units |
|  | RES4 | Temporary lodging (e.g., hotel/motel) |
|  | RES5 | Institutional dormitory (e.g., military, college, jails) |
|  | RES6 | Nursing home |
| **Commercial** | COM1 | Retail trade (e.g., stores) |
|  | COM2 | Wholesale trade (e.g., warehouses) |
|  | COM3 | Personal and repair services (e.g., service station/shop) |
|  | COM4 | Professional/technical services (e.g., offices) |
|  | COM5 | Banks |
|  | COM6 | Hospital |
|  | COM7 | Medical office/clinic |
|  | COM8 | Entertainment and recreation (e.g., restaurants/bars) |
|  | COM9 | Theaters |
|  | COM10 | Parking (e.g., garages) |
| **Industrial** | IND1 | Heavy industry (e.g., factory) |
|  | IND2 | Light industry (e.g., factory) |
|  | IND3 | Food/drugs/chemicals (e.g., factory) |
|  | IND4 | Metals/minerals processing (e.g., factory) |
|  | IND5 | High technology (e.g., factory) |
|  | IND6 | Construction (e.g., office) |
| **Agricultural** | AGR1 | Agriculture |
| **Religious** | REL1 | Church/non-profit |
| **Governmental** | GOV1 | General services (e.g., office) |
|  | GOV2 | Emergency response (e.g., police/fire station/eoc) |
| **Educational** | EDU1 | Grade schools |
|  | EDU2 | Colleges/universities (does not include group housing) |

**Table 2.** Equation and corresponding parameter values associated with building use types. See [7] for a more detailed discussion of the building use types taken from HAZUS.

| Building use type <sup>(a)</sup> | Building airflow type | R airflow parameters<br>($\lambda_{inf}$, $F_{r, fan}$, $f_{fan}$) <sup>(b)</sup> | H airflow parameters<br>($v_{fan}$, $\lambda_T$, $F_{oa}$) <sup>(c)</sup> | Deposition adjustment factor <sup>(d)</sup> | Filtration category <sup>(e)</sup> |
| --- | --- | --- | --- | --- | --- |
| Outdoors <sup>(f)</sup> | N/A | N/A | N/A | N/A | N/A |
| RES1 | R | SFH | N/A | 1 | Single family |
| RES2 | R | MH | N/A | 1.2 | Single family |
| RES3A | R | SFH | N/A | 1 | Single family |
| RES3B | R, H | APT w/o corridors | Apt w corridors | 1.2 | Low quality |
| RES3C | R, H | APT w/o corridors | Apt w corridors | 1.2 | Low quality |
| RES3D | R, H | APT w/o corridors | Apt w corridors | 1.2 | Low quality |
| RES3E | R, H | APT w/o corridors | Apt w corridors | 1 | Low quality |
| RES3F | R, H | APT w/o corridors | Apt w corridors | 1 | Low quality |
| RES4 | H | N/A | Hotel | 1 | Low quality |
| RES5 | H | N/A | Hotel | 1 | Low quality |
| RES6 | H | N/A | Hotel | 1 | Low quality |
| COM1 | H | N/A | Retail | 1 | Medium quality |
| COM2 | H | N/A | Warehouse | 0.6 | Low quality |
| COM3 | H | N/A | Retail | 1 | Low quality |
| COM4 | H | N/A | Office | 1 | Standard office |
| COM5 | H | N/A | Office | 1 | Standard office |
| COM6 | H | N/A | Health Care | 1 | Very high quality |
| COM7 | H | N/A | Health Care | 1 | Standard office |
| COM8 | H | N/A | Restaurant | 1 | Medium quality |
| COM9 | H | N/A | Retail | 1 | Medium quality |
| COM10 <sup>(g)</sup> | N/A | N/A | N/A | N/A | N/A |
| IND1 | H | N/A | Warehouse | 0.6 | Low quality |
| IND2 | H | N/A | Warehouse | 0.6 | Low quality |
| IND3 | H | N/A | Warehouse | 0.6 | High quality |
| IND4 | H | N/A | Warehouse | 0.6 | Low quality |
| IND5 | H | N/A | Warehouse | 0.6 | Very high quality |
| IND6 | H | N/A | Warehouse | 0.6 | Low quality |
| AGR | H | N/A | Warehouse | 0.6 | Low quality |
| REL1 | H | N/A | Retail | 1 | Low quality |
| GOV1 | H | N/A | Office | 1 | Standard office |
| GOV2 | H | N/A | Office | 1 | Medium quality |
| EDU1 | H | N/A | School | 1 | Medium quality |
| EDU2 | H | N/A | School | 1 | Medium quality |

<sup>(a)</sup> HAZUS building occupancy type; <sup>(b)</sup> **R** = filtered recirculation. Values selected from the distributions defined in **Table 3**; <sup>(c)</sup> **H** = HVAC system. Values selected from the distributions defined in **Tables 3 to 6**; <sup>(d)</sup> See discussion in text below; <sup>(e)</sup> Values selected from the distributions defined in **Tables 9 and 10**; <sup>(f)</sup> In this analysis, the outdoor protection factor and exit fraction is defined to be one (1); <sup>(g)</sup> Not included in the present analysis.

### 4.2. Air Flow Parameters (λ_inf_, F_r,fan_, r_fan_, v_fan_, λ_T_, F_oa_)

There are two mechanisms by which indoor and outdoor air is exchanged – via (largely) uncontrolled infiltration and via controlled mechanical means (typically HVAC or exhaust fan systems). For the single family residences, manufactured homes, duplexes, and apartments without corridors (the RES1, RES2, RES3A, and a portion of the RES3B to RES3F HAZUS building use types), **Table 3** provides the natural air infiltration rate (λ_*inf*_), furnace recirculation fan duty cycle (*F_r,fan_*), and furnace system recirculation rate (*r_fan_*) appropriate for scenarios in which windows and doors are closed. The reported geometric standard deviation accounts for variation in homes across the US and for the different seasons and times of the day.

**Table 3.** Select parameters used in R type airflow equations (filtered recirculation). These parameters assume a log-normal distribution.

| Parameter | Geometric mean<br>(units) | Geometric standard<br>deviation<br>(dimensionless) | Reference |
| --- | --- | --- | --- |
| $\lambda_{inf}$ – SFH <sup>(a)</sup> | 0.44 (h<sup>-1</sup>) | 2.04 | [14], [15] |
| $\lambda_{inf}$ – MH <sup>(a)</sup> | 0.42 (h<sup>-1</sup>) | 1.86 | [14] |
| $\lambda_{inf}$ – APT w/o corridors <sup>(a)</sup> | 0.23 (h<sup>-1</sup>) | 1.82 | [14] |
| $r_{fan}$ | 5.7 (h<sup>-1</sup>) | 1.26 | [15] |
| $F_{r, fan}$<br>(baseline scenario) | 0.25<br>(dimensionless) | 1.85 | [15] |
| $F_{r, fan}$<br>(Min MERV 7 scenario) | 1<br>(dimensionless) | 1.0 | N/A |
| $F_{r, fan}$<br>(Min MERV 11 scenario) | 1<br>(dimensionless) | 1.0 | N/A |
| $F_{r, fan}$<br>(Min MERV 14 scenario) | 1<br>(dimensionless) | 1.0 | N/A |

<sup>(a)</sup> This parameter value assumes doors and windows are closed.
SFH = single family home (used for RES1 and RES3A)
MH = manufactured home (used for RES2)
APT w/o corridors = multifamily dwelling buildings that do NOT have corridors (RES3B to RES3F w/o corridors)

**Table 4** provides our estimates for the average, minimum, and maximum values for the λ_*inf*_, *v_fan_, F_oa_*, and λ_*T*_ parameters for 9 representative US building types or subtypes: Restaurants, Offices, Schools, Retail, Health Care, Hotels (guest rooms and common spaces), Apartments with corridors, and Warehouses. [7] derives and discusses these values. As we lack a robust estimate for the functional form of the probability distribution, we assume a triangular distribution for each parameter.

**Table 4.** Select parameters used in H type airflow equations (active HVAC). These parameters assume a triangular distribution. See [7] for more details.

|  | average | maximum | minimum |
| --- | --- | --- | --- |
| supply fan rate, $v_{fan}$ ($\text{h}^{-1}$) | | | |
| Restaurant | 7.0 | 13 | 6.1 |
| Office | 3.8 | 25 | 1.1 |
| School | 3.1 | 11 | 2.8 |
| Retail | 3.7 | 9.1 | 2.0 |
| Health care | 5.8 | 18 | 3.9 |
| Warehouse | 0.9 | 1.0 | 0.6 |
| Hotel, guest rooms | 1.0 | 1.4 | 0.6 |
| Hotel, common spaces | 4.2 | 6.6 | 1.9 |
| Apt w corridors | 7.4 | 7.6 | 7.2 |
| fraction of outside air entering the building, $F_{oa}$ (dimensionless) | | | |
| Restaurant | 0.5 | 0.7 | 0.0 |
| Office | 0.1 | 1.0 | 0.0 |
| School | 0.2 | 0.6 | 0.1 |
| Retail | 0.1 | 0.6 | 0.0 |
| Health care | 0.1 | 0.2 | 0.1 |
| Warehouse | 0.05 | 0.06 | 0.04 |
| Hotel, guest rooms | 1.0 | 1.0 | 1.0 |
| Hotel, common spaces | 0.13 | 0.17 | 0.09 |
| Apt w corridors | 0.05 | 0.08 | 0.02 |
| natural air infiltration rate, $\lambda_{inf}$ ($\text{h}^{-1}$) | | | |
| Restaurant | 0.5 | 1.9 | 0.01 |
| Office | 0.12 | 1.2 | 0.0 |
| School | 0.3 | 1.2 | 0.02 |
| Retail | 0.2 | 0.8 | 0.0 |
| Health care | 0.05 | 0.9 | 0.0 |
| Warehouse | 0.3 | 1.0 | 0.05 |
| Hotel, guest rooms | 0.0 | 0.0 | 0.0 |
| Hotel, common spaces <sup>(a)</sup> | 0.3 | 1.2 | 0.0 |
| Apt w corridors | $\lambda_T - v_{fan} \cdot F_{oa}$ (see Table 5 for $\lambda_T$) | | |

<sup>(a)</sup> Assumes infiltration rates are the same for the (i) other rooms and (ii) whole building

For each of the given HAZUS apartment building use types (RES3B to RES3F), there is a mixture of buildings with and without internal corridors. We estimate the key building parameter metrics by combining both building types using the building height as a proxy for the presence of a corridor: apartment buildings greater than 3 stories are assumed to have interior corridors and apartment buildings less than 3 stories do not have them (3 story buildings were not included in the reference dataset) [14]. For the latter, we use the **R** equations as described in the prior paragraph. For the former, we use the **H** equations to incorporate a mechanical means of supplying air to the internal corridors, following *Persily et al*. [16]. **Table 5** gives the percentile distributions for the total building air ventilation rates for apartments that do have corridors. **Table 6** summarizes the fraction of buildings in each category by HAZUS building type.

**Table 5.** Percentile distribution for the total building air ventilation rate, *λ_T_* (h^−1^) for Apartments that DO have Corridors (RES3B-F) from [14].

| P1% | P5% | P25% | P50% | P75% | P95% | P99% |
| --- | --- | --- | --- | --- | --- | --- |
| 0.23 | 0.33 | 0.42 | 0.46 | 0.54 | 0.71 | 0.87 |

**Table 6.** Fraction of apartment buildings by HAZUS building use type that are either less than or greater than 3 stories in height (from [16]).

| HAZUS building use type | HAZUS building use type description | Fraction of buildings less than 3 stories (dimensionless) | Fraction of buildings greater than 3 stories (dimensionless) |
| --- | --- | --- | --- |
| RES3B<sup>(a)</sup> | Multi-family dwelling: 3–4 units | 0.87 | 0.13 |
| RES3C | Multi-family dwelling: 5–9 units | 0.80 | 0.20 |
| RES3D | Multi-family dwelling: 10–19 units | 0.60 | 0.40 |
| RES3E<sup>(b)</sup> | Multi-family dwelling: 20–49 units | 0.24 | 0.76 |
| RES3F<sup>(c)</sup> | Multi-family dwelling: 50+ units | 0.10 | 0.90 |

<sup>(a)</sup> Persily et al. [16] results correspond to buildings with 2 to 4 units.
<sup>(b)</sup> Persily et al. [16] results correspond to buildings with 20 to 39 units.
<sup>(c)</sup> Persily et al. [16] results correspond to buildings with 40+ units.

Hotels (RES4), and presumably other institutional residential buildings such as dormitories and nursing homes (RES5 and RES6), can have different air handling mechanisms in the common use spaces (e.g., lobby, restaurants/dining areas, corridors) compared to the sleeping areas (e.g., guest rooms). As discussed in [7], there is minimal mixing between these two regions, and so we develop two, independent sets of building protection estimates – one for guest rooms and the other for the rest of the building, see **Table 4**. We note that the population in these two building areas varies by the time of day – with nearly all people in the guest room at night and more evenly distributed throughout the building during the day [17]. As a result, the appropriate building protection factors vary by time of day.

### 4.3. Particle Size Specific Parameters (λ_dep_, L_inf_, F_filter_)

Indoor particle concentrations are affected by three parameters: deposition to indoor surfaces, losses that occur when airborne particles penetrate through the building envelope, and losses that occur during filtration. Particle deposition loss rates are primarily controlled by particle size, the indoor surface to volume ratio, and, to some extent, turbulence conditions within a given space. We treat airborne particles as chemically inert, with negligible thermophoretic or electrostatic interactions with indoor surfaces in buildings. **Table 7** provides the particle deposition loss rate distributions for residential buildings and is based on experiments conducted in actual homes or room-sized chambers. There are almost no comparable, size-resolved particle deposition rate data reported for other building types. For these buildings, we have used the residential loss rates and, for some cases, multiplied the residential loss rates by our estimated deposition adjustment factor. This factor is based on our estimates of the differences in surface-to-volume ratios in those buildings compared with those in residences and accounts for the quantity and type of furnishings. These factors are shown in the next to the last column of **Table 2**.

**Table 7.** Cumulative frequency distribution of airborne particle deposition loss rates in buildings (λ_dep_, h^−1^) by particle size (aerodynamic diameter). See [7] for more details.

| Airborne particle size ($\mu\text{m}$) | Percentile distribution for the indoor deposition particle loss rate ($\lambda_{\text{dep}}$, $\text{h}^{-1}$) | | | | | | |
| --- | --- | --- | --- | --- | --- | --- | --- |
|  | P<sub>1%</sub> | P<sub>5%</sub> | P<sub>25%</sub> | P<sub>50%</sub> | P<sub>75%</sub> | P<sub>95%</sub> | P<sub>99%</sub> |
| 0.1 | 0.07 | 0.07 | 0.39 | 0.59 | 0.72 | 1.57 | 1.67 |
| 0.3 | 0.02 | 0.11 | 0.27 | 0.46 | 0.93 | 1.31 | 1.35 |
| 1 | 0.04 | 0.15 | 0.28 | 0.40 | 0.89 | 2.39 | 2.68 |
| 3 | 0.05 | 0.31 | 0.72 | 1.31 | 1.87 | 3.60 | 3.89 |
| 10 | 0.08 | 0.08 | 1.83 | 4.12 | 6.78 | 10.83 | 11.45 |

Airflow pathways into buildings depend upon a number of building features, primary among them is whether the building has an HVAC system, as most commercial buildings do. Residential buildings, on the other hand, largely do not and thus the major airflow pathway is infiltration through cracks and penetrations in the building shell. As discussed in greater detail in [7], particle losses are parameterized by the particle penetration efficiency, which is particle-size-dependent; values for these parameters are presented in **Table 8** and are used here for all building types.

**Table 8.**
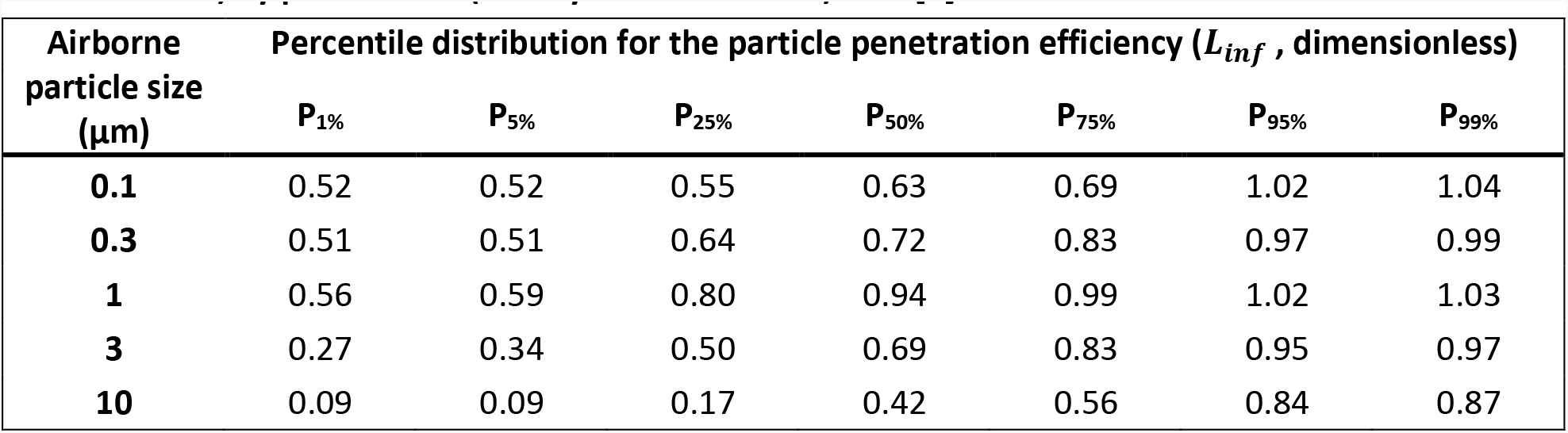
Cumulative frequency distribution of the building particle penetration efficiency (L_inf_, dimensionless) by particle size (aerodynamic diameter). See [7] for more details.

For buildings with HVAC systems, a major pathway for outdoor air entering the building is through the mechanical air handling system which typically contains some particle filtration capability. As discussed in [7], the performance of these particle filters is typically categorized by their Minimum Efficiency Reporting Value (MERV) rating, as specified by ASHRAE [18]. MERV ratings correspond to the fraction of airborne particles captured in a single pass through the filter. Filters with numerically larger MERV ratings are more efficient at removing airborne particles than filters with numerically smaller MERV ratings. For our purposes here, we consider 6 filtration categories – single family, standard office, low quality, medium quality, high quality, and very high quality – each with a different distribution of filter MERV ratings. For our baseline scenario calculation purposes, we assign each HAZUS building use type the filtration category corresponding to our estimated distribution of the filter media types and quality that are in general use (see last column in **Table 2**).

As we are analyzing the effects of improved filtration efficiency, the filtration category specific distribution of MERV rated filters varies for each scenario analyzed as shown in **Table 9**. The *baseline scenario* distribution is compatible with prior literature surveys of US building filtration efficiency and is discussed in [7]. The other three scenarios assume that the currently existing lower efficiency furnace and HVAC filters are replaced with progressively higher efficiency filters. The difference between the latter three scenarios is the minimum filter rating considered. The *Minimum MERV 7 scenario* replaces all filters with MERV ratings below MERV 7 with a MERV 7 or 8 filter (other filters are not changed). The *Minimum MERV 11 scenario* replaces all filters with a MERV rating below 11 with a MERV 11 or 12 filter (other filters are not changed). The *Minimum MERV 14 scenario* replaces all filters with a MERV rating below 14 with a MERV 14 or 15 rated filter. We note that 35% of single family homes do not have recirculating air furnaces – e.g., radiators, in-wall heaters – and so their building filtration remains unchanged in all scenarios. The computed results for each filtration scenario include these homes.

For each MERV filter rating shown in **Table 9, Table 10** provides the single pass filtration efficiency distributions which, for each MERV rating, considers the efficiency variation both (a) within similarly rated filters and (b) due to filter loading over the filter lifetime (note, our filtration parameters are for media-type filters). See [7] for more details.

**Table 9.**
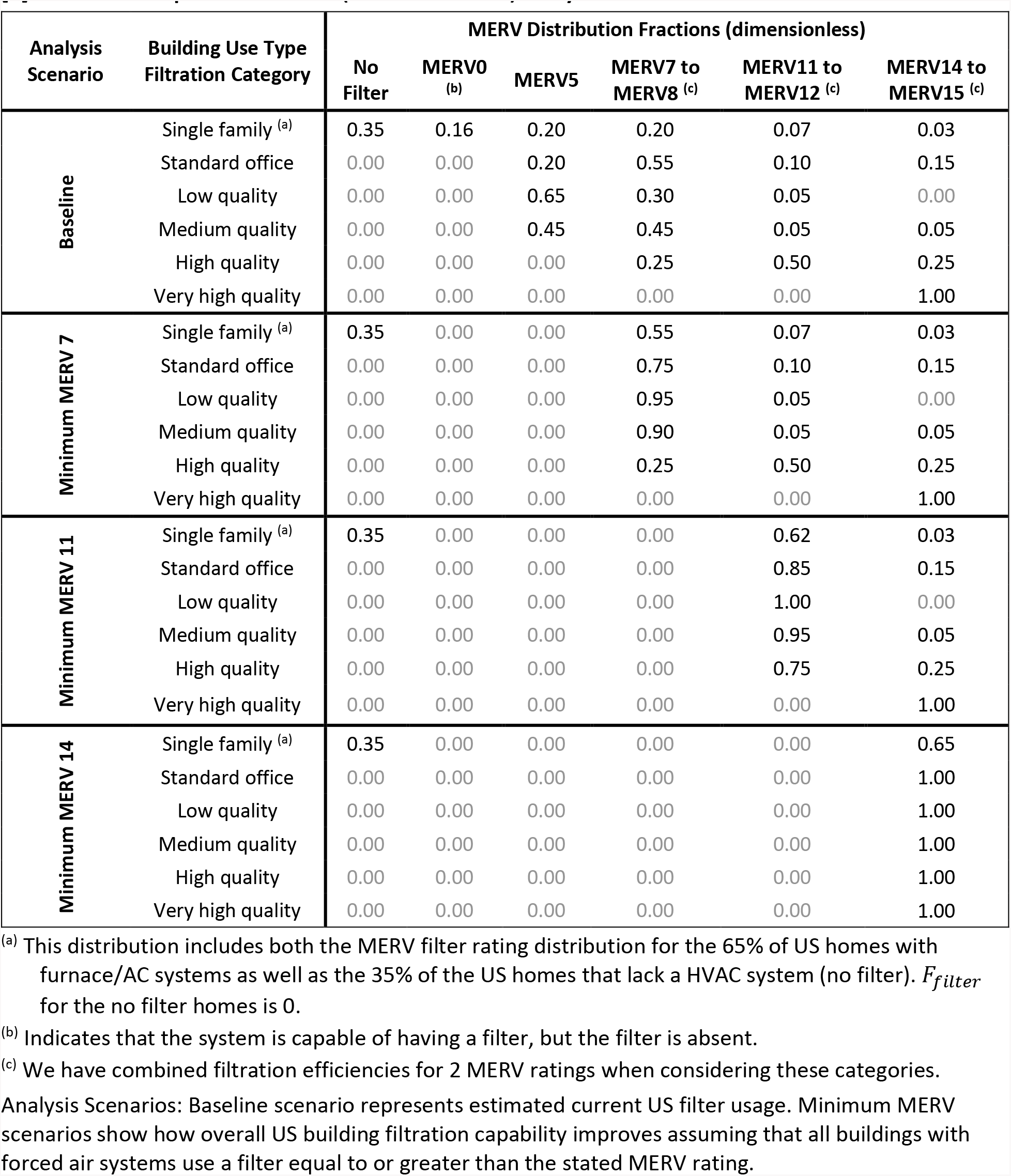
Distributions of MERV-rated filters by building use type filtration categories for the baseline [7] and for the improved filtration (Minimum MERV) analysis scenarios.

**Table 10.** Filtration efficiency (F_filter_, dimensionless) by air filter MERV rating and particle size. See [7] for more details.

| Air-filter<br>MERV<br>rating | Airborne<br>particle size<br>($\mu\text{m}$) | Percentile distribution for the filtration efficiency ($F_{filter}$, dimensionless) | | | | | | |
| --- | --- | --- | --- | --- | --- | --- | --- | --- |
|  |  | P<sub>1%</sub> | P<sub>5%</sub> | P<sub>25%</sub> | P<sub>50%</sub> | P<sub>75%</sub> | P<sub>95%</sub> | P<sub>99%</sub> |
| **0** | 0.1 | 0 | 0 | 0 | 0 | 0 | 0 | 0 |
|  | 0.3 | 0 | 0 | 0 | 0 | 0 | 0 | 0 |
|  | 1 | 0 | 0 | 0 | 0 | 0 | 0 | 0 |
|  | 3 | 0 | 0 | 0 | 0 | 0 | 0 | 0 |
|  | 10 | 0 | 0 | 0 | 0 | 0 | 0 | 0 |
| **5** | 0.1 | 0.00 | 0.00 | 0.00 | 0.01 | 0.04 | 0.18 | 0.29 |
|  | 0.3 | 0.00 | 0.00 | 0.00 | 0.01 | 0.04 | 0.18 | 0.29 |
|  | 1 | 0.05 | 0.05 | 0.07 | 0.10 | 0.18 | 0.69 | 0.83 |
|  | 3 | 0.28 | 0.28 | 0.32 | 0.40 | 0.64 | 0.95 | 0.98 |
|  | 10 | 0.22 | 0.22 | 0.29 | 0.45 | 0.83 | 0.94 | 0.98 |
| **7 to 8** | 0.1 | 0.03 | 0.03 | 0.12 | 0.20 | 0.24 | 0.33 | 0.37 |
|  | 0.3 | 0.03 | 0.03 | 0.12 | 0.20 | 0.24 | 0.33 | 0.37 |
|  | 1 | 0.15 | 0.27 | 0.51 | 0.69 | 0.81 | 0.90 | 0.92 |
|  | 3 | 0.51 | 0.58 | 0.93 | 0.96 | 0.98 | 0.99 | 1.00 |
|  | 10 | 0.61 | 0.62 | 0.95 | 0.99 | 1.00 | 1.00 | 1.00 |
| **11 to 12** | 0.1 | 0.04 | 0.04 | 0.07 | 0.40 | 0.56 | 0.92 | 0.94 |
|  | 0.3 | 0.04 | 0.04 | 0.07 | 0.40 | 0.56 | 0.92 | 0.94 |
|  | 1 | 0.22 | 0.25 | 0.42 | 0.76 | 0.91 | 0.99 | 0.99 |
|  | 3 | 0.67 | 0.68 | 0.87 | 0.95 | 0.99 | 1.00 | 1.00 |
|  | 10 | 0.67 | 0.67 | 0.85 | 0.98 | 1.00 | 1.00 | 1.00 |
| **14 to 15** | 0.1 | 0.64 | 0.68 | 0.80 | 0.86 | 0.92 | 0.99 | 0.99 |
|  | 0.3 | 0.64 | 0.68 | 0.80 | 0.86 | 0.92 | 0.99 | 0.99 |
|  | 1 | 0.86 | 0.90 | 0.96 | 0.98 | 0.99 | 1.00 | 1.00 |
|  | 3 | 0.98 | 0.99 | 0.99 | 1.00 | 1.00 | 1.00 | 1.00 |
|  | 10 | 0.99 | 0.99 | 1.00 | 1.00 | 1.00 | 1.00 | 1.00 |

## 5. Results

Using the equations, input assumptions, and data described in the previous two sections, we estimate the benefits of improving building filtration efficiency using off-the-shelf air filter technology across 32 of the 33 major FEMA HAZUS building use types shown in **Table 1**. In this section we present these benefits by comparing three improved filtration scenarios, each with increasingly higher minimum filtration efficiency, against a baseline scenario, which reflects our understanding of the existing filtration distribution in the US building stock and normal furnace operating conditions. Three primary metrics are considered here: (1) the passive protection provided by buildings against outdoor airborne particulate hazards (Building Transmission Factor), (2) the degree to which indoor individuals are exposed to indoor-origin airborne particles (Indoor Normalized TSIAC) and (3) the fraction of indoor airborne particles that exit the building and enter the outdoor atmosphere (Building Exit Fraction). We also discuss the potential improvement on regional exposures due to particles exiting the building and exposing downwind building occupants. For each of the 32 building use types, our modeling covers five aerosol sizes (0.1, 0.3, 1, 3, 10 µm diameter) and four airborne loss rates (0, 0.1, 1, 10 hr^−1^). The entire set of results are provided in **Supplemental Material A: Detailed Results**.

We focus the discussion of these results on six common building types, two aerosols sizes (1 µm and 3 µm aerodynamic diameter), an airborne aerosol decay rate of 0 hr^−1^, and the US average (expectation value) result for each building metric. The six building types consist of three residential building categories: single family houses (RES1), small apartment buildings with 3 to 9 units (average of RES3B and RES3C), and large apartment buildings with 20 to 50+ units (average of RES3E and RES3F). It is important to note that the modeling results for the two types of apartment buildings are based on combinations of buildings with different air handling systems. The three non-residential building categories are: retail stores (COM1), office buildings (average of COM4, COM5 and GOV1), and schools (EDU1).

Results are presented graphically for the baseline scenario and then as the fractional improvements resulting from better filtration. Summary tables are provided in **Supplemental Material B: Results Summary**. Additional figures showing the improved filtration scenario results are provided in **Supplemental Material C: Additional Building Metric Figures**. Finally, the systematic influence of particle size and airborne decay rate on the improvement ratio for the Building Transmission Factor is illustrated for MERV 14–15 filters in **Supplemental Material D: Improvement Sensitivity to Particle Size and Airborne Loss Rate**.

A formal uncertainty analysis was not performed for the metrics presented in this section. For each metric and filter scenario we have presented the average results from ∼10,000 Monte Carlo modeling runs because average values are commonly used in air quality exposure and health impact assessments. The corresponding modeling uncertainty in the mean value, the standard error, is a few percent. In contrast, robust estimates of the input parameter uncertainty are not available, see [7] for more detail, and so result uncertainty due to input parameter uncertainty was not generated. The model outputs shown in **Supplemental Material A: Detailed Results** are presented as the averages of five equally sized bins into which the model results are aggregated. These bins range from the lowest 20% of the model results to the highest 20% and so provide an estimate of the overall result variability. In most cases, this variation is within a factor of 1.5 to 4 of the average value.

### 5.1. Reducing Indoor Exposures to Outdoor Airborne Particles (Building Transmission Factor)

The baseline Building Transmission Factor results, shown in **Figure 1**, reflect our estimates of the ratio of the indoor exposures to the corresponding outdoor exposure for the existing filtration distribution and normal furnace operating conditions for the 6 common US building types we have chosen to highlight. There is a wide range of Building Transmission Factor estimates, with the best protection (lowest ratios) for apartments. For most small apartment buildings, infiltration rates are small, and so the source term (numerator) in **Equation 3R** is small, see **Table 3**. For most large apartments buildings, our analysis assumes an active HVAC system (**Equation 3H**), with a relatively larger recirculation rate, see **Table 4**. In general, buildings provide more protection against 3 µm diameter aerosols, as compared to 1 µm diameter aerosols, mainly due to higher filtration efficiencies for the larger particles – even for lower efficiency filters – and higher indoor deposition loss rates.

**Figure 1.**
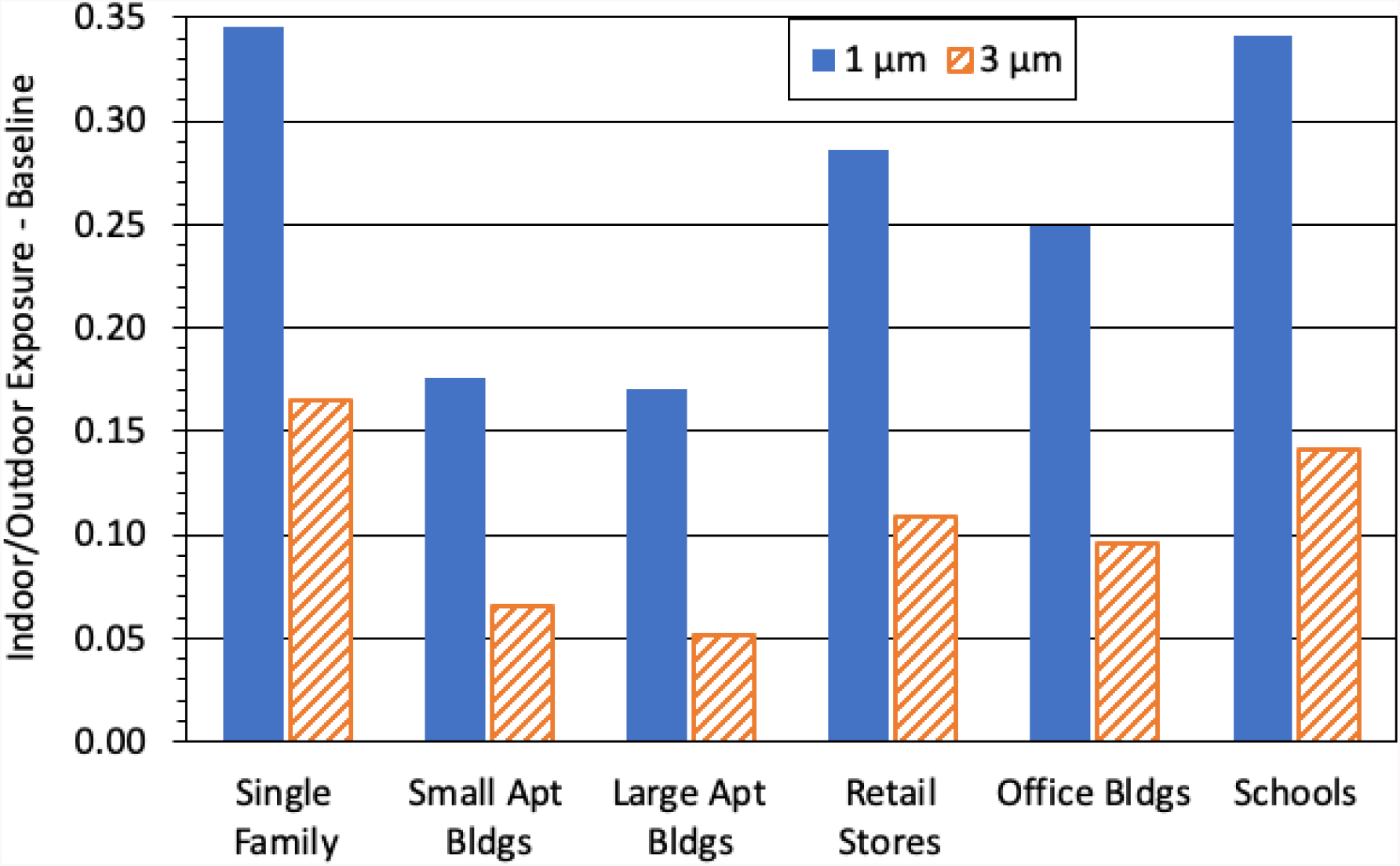
Baseline scenario results for the US Building Transmission Factors (dimensionless) – the ratio of the indoor exposures to the corresponding outdoor exposure – for selected building types; and two aerosol sizes: 1 µm (solid bar) and 3 µm diameter (diagonal striped bar). Higher Building Transmission Factor values indicate more exposure.

Figure 2 presents the improvement associated with higher efficiency filtration using the ratio of Building Transmission Factor for the baseline scenario compared to the specified improved filtration scenario for selected residential and non-residential buildings. A value of 1 on the vertical axis indicates no improvement over the baseline scenario. A factor of 2 means that the improved filtration scenario results in half the Building Transmission Factor (and thus half the indoor exposure) relative to the baseline scenario. Recall that improved filtration scenarios assume the furnace fan duty cycle is increased to 100% (always on) in those buildings with forced air heating/cooling systems. **Figure 2** shows that for all building types, scenarios using a MERV 7 or greater filter provide at least a 35% reduction in indoor exposures for 1 µm and 3 µm diameter particles. The reductions in indoor exposures vary by building type, minimum MERV filter rating, and particle size.

**Figure 2.**
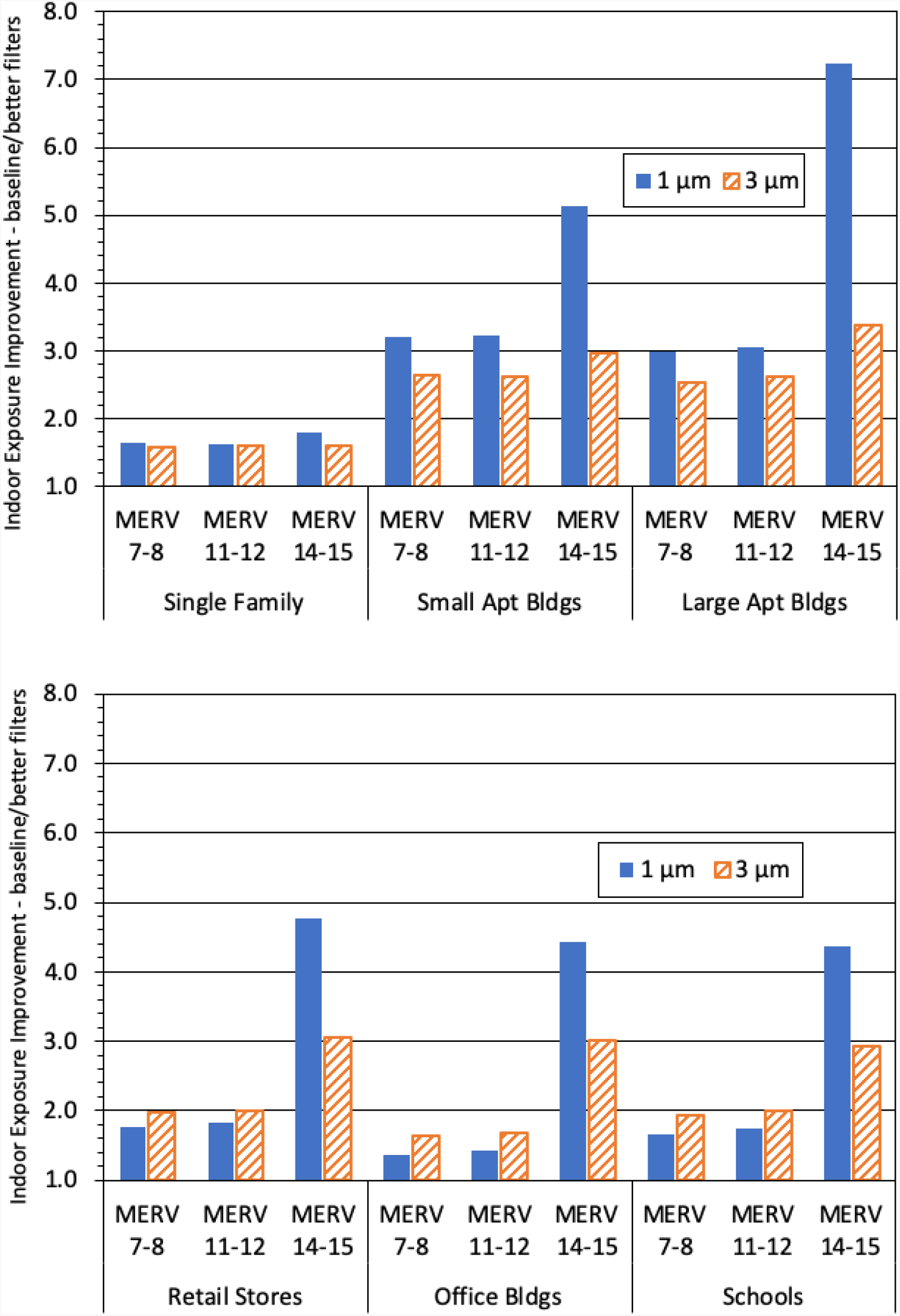
Improvement in the US Building Transmission Factor for selected residential (top panel) and non-residential (bottom panel) for selected building types; two aerosol sizes: 1 µm (solid bar) and 3 µm diameter (diagonal striped bar); and 3 minimum MERV ratings. The vertical axis is the ratio of the Building Transmission Factors for the baseline to the specified improved filtration scenario (a value of 1 indicates no improvement, larger numbers indicate greater improvement).

Figure 2 (top panel) shows that for single family dwellings, the progression across the improved filtration scenarios yields a factor of 1.6x to 1.8x improvement for the two particle sizes shown (i.e., ∼40% reduction in indoor exposures). For apartments, the magnitude of the improvement is larger, a factor of 2.5x to 3.2x (∼65% reduction) for the Minimum MERV 7 and Minimum MERV 11 scenarios. For large apartments, an even greater improvement occurs with the use of high efficiency MERV 14 or 15 rated filters, e.g., a factor of 3.5x to 7x improvement (∼80% reduction). The larger improvement for apartments compared with single family residences is due, in part, to four factors. First, 35% of the modeled single family residences do not have forced air systems, and so improved filtration is possible only for 65% of these buildings. Second, 65% of the apartment filters in the baseline scenario have low particle filtration efficiency (MERV 5). Third, most smaller apartment buildings have lower infiltration rates. Fourth, most larger apartment buildings rapidly recirculate indoor air through a filter contained within the local heating/cooling system.

Figure 2 (bottom panel) shows the corresponding improvement for the three selected non-residential building types. Overall, the results show patterns similar to those discussed for the residential building types where the improvement for the Minimum MERV 7 and Minimum MERV 11 scenarios are similar, a factor of 1.7x to 2x improvement (∼45% reduction) for retail stores and schools and somewhat lower, a factor of 1.4x to 1.7x improvement (∼35% reduction), for office buildings. Again for most building types, notably greater improvement occurs with the use of high efficiency MERV 14 or 15 rated filters, i.e., a factor of 3x to 5x improvement (∼70% reduction), and this effect is most noticeable for 1 µm particles.

### 5.2. Reducing Indoor Exposures to Indoor Airborne Particles (Indoor Normalized TSIAC)

The baseline Indoor Normalized TSIAC results, shown in **Figure 3**, reflect our estimates of the indoor exposure to airborne particles released indoors for six common US building types. There is a wide range of Indoor Normalized TSIAC estimates, with the highest exposures (higher values) present in single family homes and small apartment buildings and the lowest exposures in office buildings. Exposures are lower for 3 µm diameter aerosols, as compared to 1 µm diameter aerosols. These results indicate that for particles released indoors, higher ventilation rates, combined with better particle filtration present in office and school buildings, provides lower exposures in non-residential buildings than is the case in residential buildings with lower ventilation rates and reduced filtration capability.

**Figure 3.**
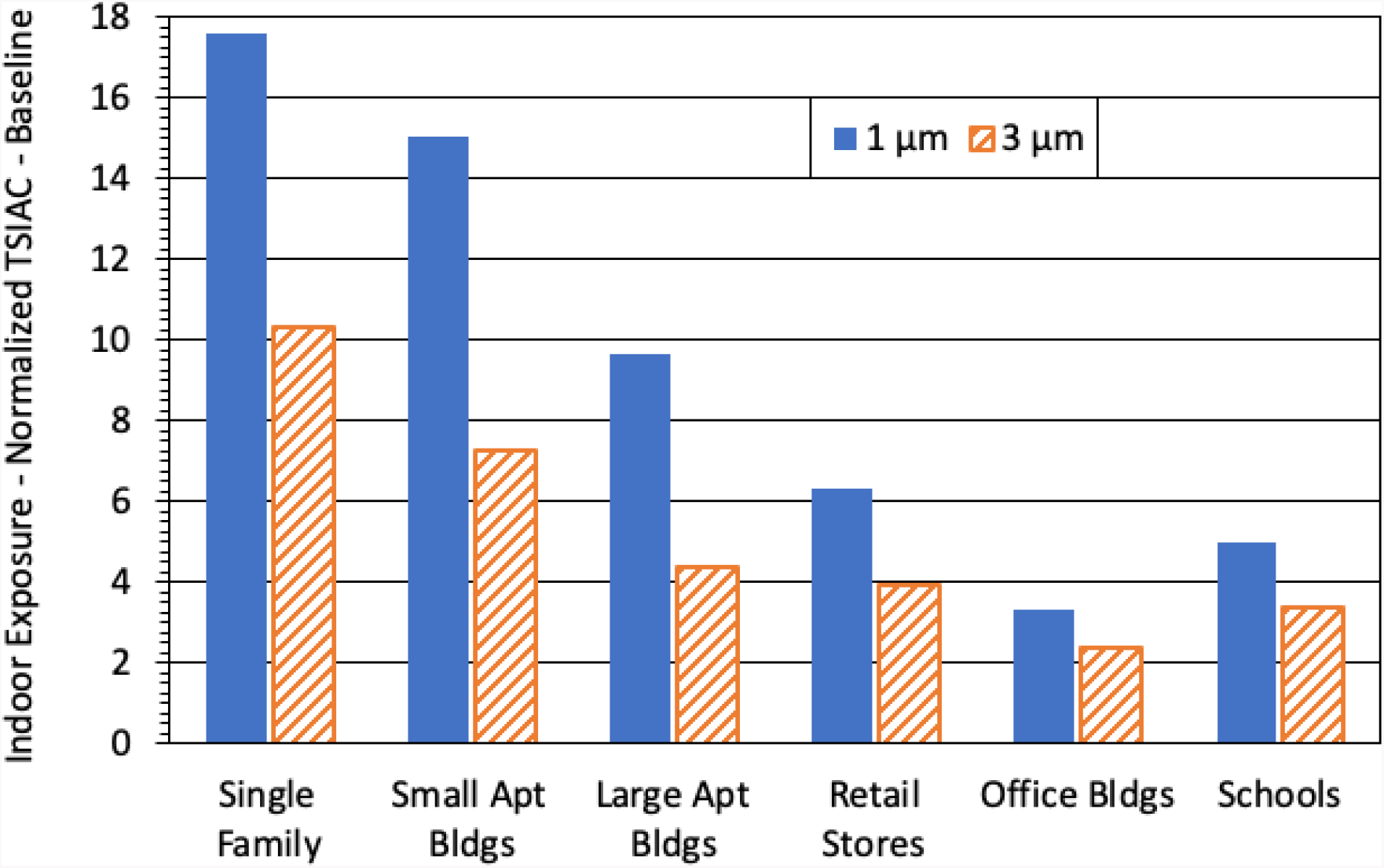
Baseline scenario results for the US Indoor Normalized Time and Space Integrated Air Concentration (Indoor Normalized TSIAC in s m^−1^) – indoor exposures to indoor-origin airborne particles – for selected building types; and two aerosol sizes: 1 µm (solid bar) and 3 µm diameter (diagonal striped bar). Higher Indoor Normalized TSIAC values indicate more exposure.

Figure 4 presents the improvements associated with higher efficiency filtration using the ratio of the Indoor Normalized TSIAC for the baseline scenario compared to the specified improved filtration scenario for selected residential and non-residential buildings. In general, all building types using a MERV 7 or higher rated filters have reductions in indoor exposures, however the reduction is greater in residential buildings, with apartment buildings showing the largest improvements.

**Figure 4.**
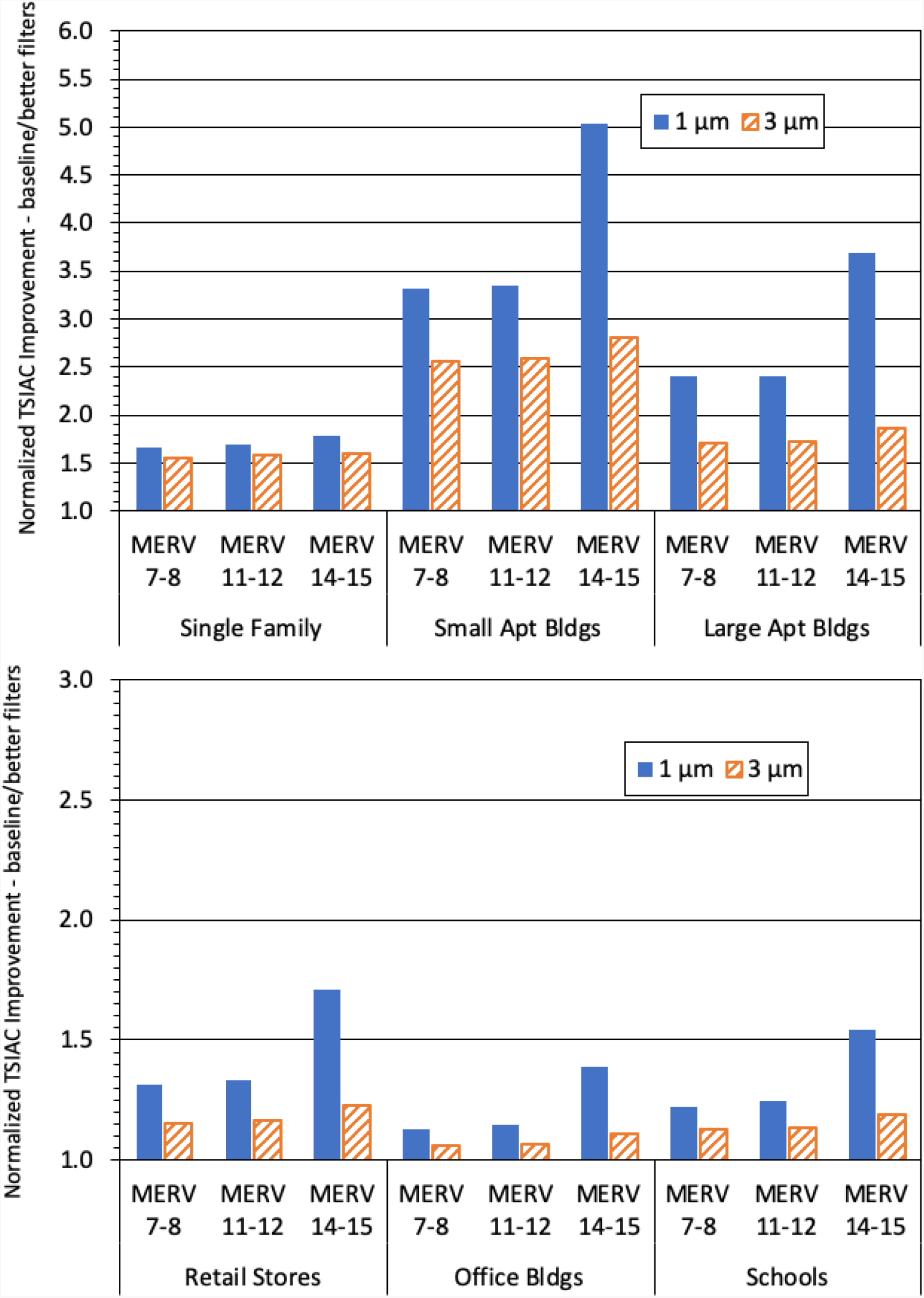
Improvement in the US Indoor Normalized TSIAC for selected residential (top panel) and non-residential (bottom panel) building types; two aerosol sizes: 1 µm (solid bar) and 3 µm diameter (diagonal striped bar); and 3 minimum MERV ratings. The vertical axis shows the ratio of the Indoor Normalized TSIAC for the baseline scenario to the specified improved filtration scenario (a value of 1 indicates no improvement, larger numbers indicate greater improvement). Note the factor of 2 difference in vertical scales between the top and bottom panels.

Figure 4 (top panel) shows that for single family dwellings, the progression across the improved filtration scenarios yields a factor of 1.5x to 1.8x improvement for the two particle sizes shown (∼40% reduction in indoor exposures). For apartments, the magnitude of the improvement is larger, a factor of 1.7× to 3.4× (∼55% reduction) for the Minimum MERV 7 and MERV 11 scenarios. Again, greater improvement occurs with the use of high efficiency MERV 14 or 15 rated filters, e.g., a factor of 1.9x to 5x improvement (45% to 80% reduction) for apartment buildings. Again, the larger improvement for apartments compared with single family residences is due, in part, to our assumption that many apartments currently have low efficiency filters, and some, often smaller, US apartment buildings have lower infiltration rates while other, often larger, apartments rapidly recirculate indoor air through a filter contained within the local heating/cooling system.

Figure 4 (bottom panel) shows the corresponding improvement for the three selected non-residential building types. Overall, the results show smaller improvements than those discussed for the residential building types, a factor of 1.1× to 1.3× improvement (∼15% exposure reduction) for the Minimum MERV 7 and Minimum MERV 11 scenarios. Greater improvement occurs with the use of high efficiency MERV 14 or 15 rated filters, e.g., a factor of 1.1× to 1.7× improvement (10% to 40% reduction). Exposures in non-residential buildings are less responsive to improved filtration because the baseline filter quality distribution for nonresidential buildings is higher and because in most non-residential buildings the HVAC system is already assumed to be operating 100% of the time.

### 5.3. Fraction of Indoor Airborne Particles Released to the Outdoors (Building Exit Fraction)

Indoor air containing airborne particles exfiltrates and/or is mechanically exhausted from buildings, providing a source of outdoor airborne particles. The baseline Building Exit Fraction estimates, shown in **Figure 5**, reflect our estimates of the fraction of indoor airborne particles that exit the selected US building types. There is a wide range of Building Exit Fraction estimates, with the highest fractions (higher values) present in offices and school buildings and the lowest in apartment buildings. Because the non-residential buildings have HVAC systems, typically with an integral exhaust pathway, these buildings release more particles to outdoors while for residential buildings, the exfiltration rate is lower and includes additional particle losses as the exfiltrating air passes through cracks and other openings in the building shell. Fewer 3 µm diameter particles, as compared to 1 µm diameter particles, are released to the outdoors due to higher within-building losses for the 3 µm diameter particles.

The indoor exposures to an indoor airborne particle (Indoor Normalized TSIAC) and the fraction of indoor, airborne particles exiting the building are closely related, see **Equation 7**. As a result, the Building Exit Fraction reductions for the improved filtration scenarios, **Figure 6**, relative to the baseline scenario, are nearly identical to, but slightly lower than, the indoor exposure improvements shown in the previous section, **Figure 4**. The difference is due to the particle losses that occur during exfiltration.

**Figure 5.**
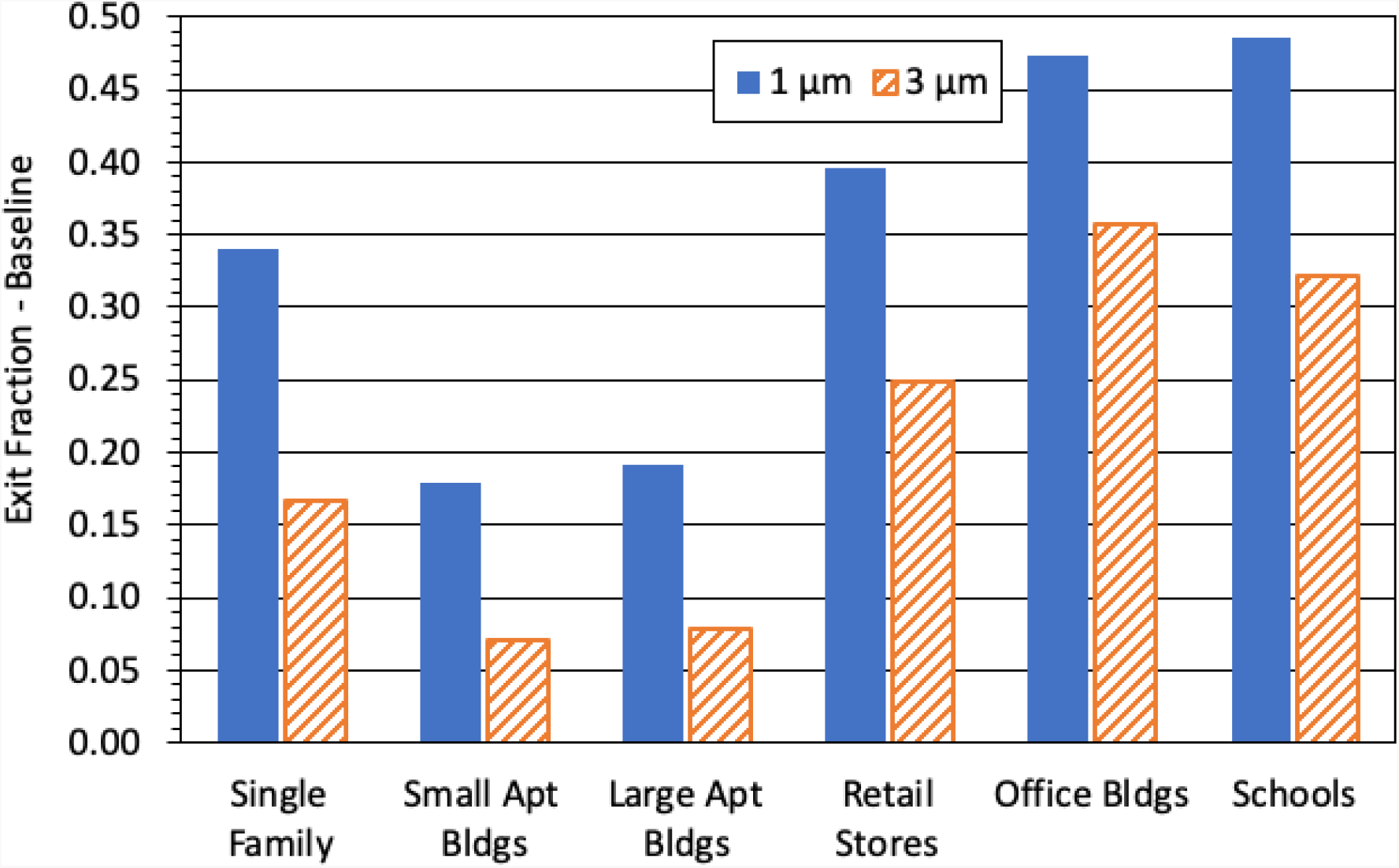
Baseline scenario results for the US Building Exit Fraction (dimensionless) – the fraction of indoor airborne particles that are released to the outdoor atmosphere – for selected building types; and two aerosol sizes: 1 µm (solid bar) and 3 µm diameter (diagonal striped bar). Higher Exit Fraction values correspond to more particles released to the outdoor atmosphere.

**Figure 6.**
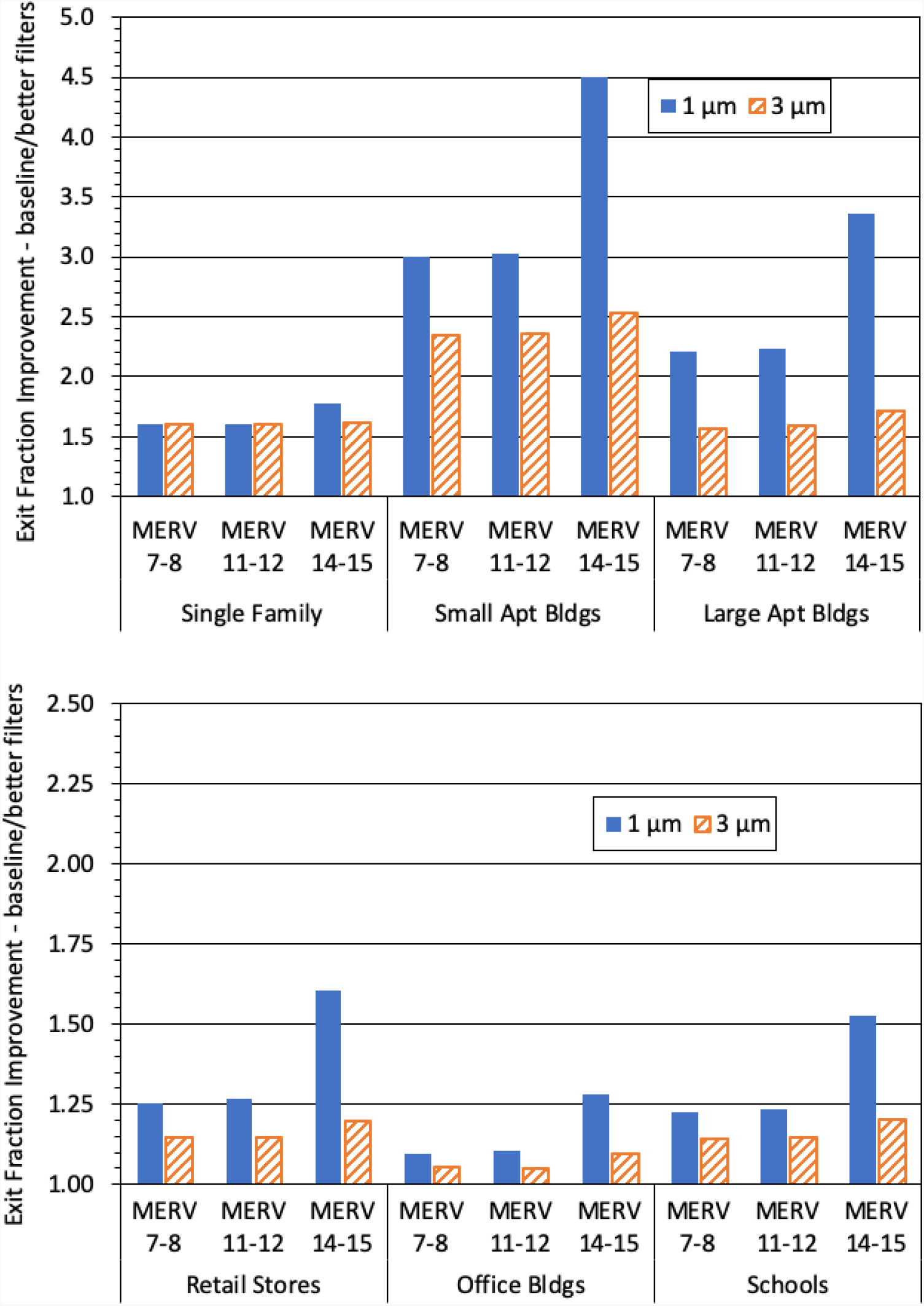
Improvement in the US Building Exit Fraction for selected residential (top panel) and non-residential (bottom panel) building types; two aerosol sizes: 1 µm (solid bar) and 3 µm diameter (diagonal striped bar); and 3 minimum MERV ratings. The vertical axis shows the ratio of Building Exit Fraction for the baseline scenario to the specified improved filtration scenario (a value of 1 indicates no improvement, larger numbers indicate greater improvement). Note the factor of 2 difference in vertical scales between the top and bottom panels.

### 5.4. Regional Exposures

Particles can exit a building, be transported downwind, and either expose outdoor individuals or enter a second building to expose indoor, downwind individuals. Improving building air filtration reduces indoor, downwind exposures through decreases in both the (a) fraction of indoor airborne particles that exit the source building (Building Exit Fraction) and (b) the fraction of outdoor airborne particles that enter the downwind building indoor air (the downwind Building Transmission Factor), see **Equation 9**.

In general particles can be emitted from, and expose downwind individuals in, a wide variety of different building types and a full Regional Shelter Analysis is required to model this common general case where emissions and exposures occur across multiple building types, e.g., [6], [7]. While a full RSA is beyond the scope of the current study, estimates for overall air filtration improvement benefits can be obtained for neighborhoods in which the population resides in a homogenous housing stock.

Figure 7 presents the improvement associated with higher efficiency filtration using the ratio of the Downwind Indoor Exposure for the baseline scenario compared to the specified improved filtration scenario for selected residential buildings, see **Equation 10**. A value of 1 on the vertical axis indicates no improvement over the baseline scenario. Larger numbers indicate greater improvement.

In neighborhoods populated with single family homes, the Minimum MERV 7 and Minimum MERV 11 scenarios provide a factor of 2.6x improvement for the two particle sizes shown (60% reduction in downwind indoor exposures). The Minimum MERV 14 scenario providing slightly more protection, a factor of 2.6x to 3.2x (65% reduction). The improvement is notably greater for neighborhoods populated with apartment buildings, with a 4x to 10x improvement (75% to 90% reduction) for the Minimum MERV 7 and Minimum MERV 11 scenarios. The improvement for the Minimum MERV 14 scenario ranges from 5.8x to 25x (85% to 96% reduction). In the above results, the improvement for larger 3 µm particles is less than that for 1 µm particles, especially so for the MERV 14 or 15 rated filters.

**Figure 7.**
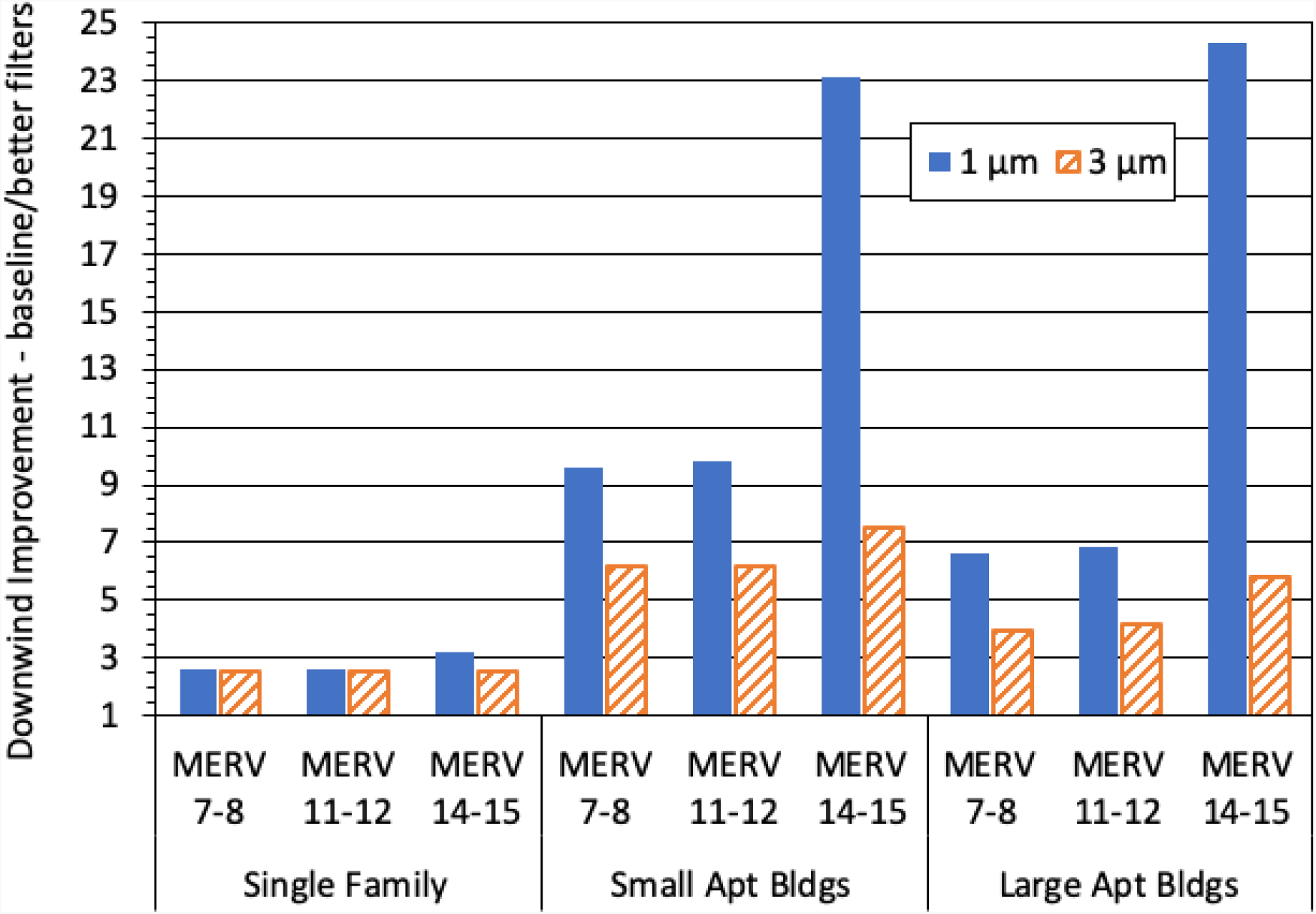
Improvement in the US Downwind Indoor Exposure for homogenous residential neighborhoods comprised of the selected building use types; two aerosol sizes: 1 µm (solid bar) and 3 µm diameter (diagonal striped bar); and 3 minimum MERV ratings. The vertical axis shows the ratio of the Downwind Indoor Exposure for the baseline scenario to the specified improved filtration scenario (a value of 1 indicates no improvement, larger numbers indicate greater improvement).

## 6. Discussion and Conclusion

This work broadly evaluates the extent to which improved airborne particle filtration in US buildings may reduce airborne particle exposures. This work is based on an aggregation of building types and a general set of parameters that describes both airflows into and out of buildings as well as indoor particle behavior and removal processes. Our analysis suggests that, for most building types studied, upgrades to the filters currently used in furnaces or HVAC systems may reduce airborne particle exposures – although the degree of improvement varies by filter efficiency, particle size, building type, and specific building. Our modeling indicates that there is limited difference between scenarios which impose a minimum filter rating of MERV 7 vs. MERV 11, although both improved filtration scenarios are expected to reduce exposures relative to the existing building stock. The largest reductions in airborne particulate exposures are associated with the use of high filtration efficiency (MERV 14 or 15 rated) filters and, in many cases, 1 µm diameter particles, reflecting their typically larger improvements in filtration efficiency for this particle size.

As a building use type, apartments benefit the most from improved, within-building filtration, in part, due to our estimate that 65% of apartments currently have low efficiency (MERV 5 rated) filters. In addition, and relative to other building use types examined, most smaller apartment buildings have lower infiltration rates while most larger apartments rapidly recirculate indoor air through a filter contained within the local heating/cooling system. Note that in order to conform to the HAZUS building use types, we have modelled these two apartment categories with a mix of heating/cooling system and air infiltration characteristics.

In contrast, the calculated improvement for single family homes is relatively modest due, in part, to the estimated 35% of single family homes have heating systems, such as radiators and in-wall heaters, that do not have filters and therefore cannot benefit from the use of improved filters. This assumption limits the overall improvement, particularly for the Minimum MERV 14 scenario. For example, the Building Transmission Factor improves by a factor of 1.8x for the Minimum MERV 14 scenario when considering the entire US building stock, but a factor of 5.8x when considering the best protected buildings with filtered air handling systems (60% of the building stock)^1^. This point is also pertinent when comparing our modeling results with other studies results as discussed below.

### 6.1. Comparison to Prior Work

While there are ample data on the filtration efficiency of various filters and filter types, there are fewer studies that have experimentally assessed the performance of filters in whole buildings. Most are research studies conducted in one or a small number of buildings. Hence the data available for comparison with our modeling results are not broadly representative of the building types studied here. We summarize below a selected (non-comprehensive) set of results from such studies to provide context for our current work. We compile the results from these studies – along with our comparative modeling results – in **Table 11**. The pertinent details of the previous studies are discussed below.

*Howard-Reed et al*. [19] performed one of the first experimental assessments of the effect of filter efficiency on indoor particle concentrations in a normally occupied home. *Howard-Reed et al*. compared the use of a ‘typical’ panel furnace filter (PFF) with that of an in-duct electrostatic precipitator (ESP). Although the authors did not provide filter ratings for these devices, the filtration efficiencies measured in their tests are approximately consistent with the MERV 5 and MERV 12 filtration efficiencies shown in **Table 10** above for the PFF and ESP, respectively. The furnace fan was operated only when particle concentration measurements were conducted, and then only following the times when particles were generated indoors. Similar tests were conducted in an unoccupied test house that had a lower indoor/outdoor air change rate than the occupied dwelling. Using the measured filtration rate and particle loss rate data, the authors estimated the effect of filtration on the ratios of the concentrations of indoor particles of outdoor origin to outdoor particle concentrations. The estimated indoor/outdoor ratios for the PFF and ESP cases are given in **Table 11** and are consistent with the range of Building Transmission Factors that we predict for single family homes.^2^ The *Howard-Reed et al*. data show a definite improvement that is somewhat larger than the improvement predicted by our modeling work for the US single family home building stock. However, the reported improvement with the better (ESP) filter is similar to our prediction that considers only the portion of the single family building stock that has filtration-capable heating systems.

**Table 11.**
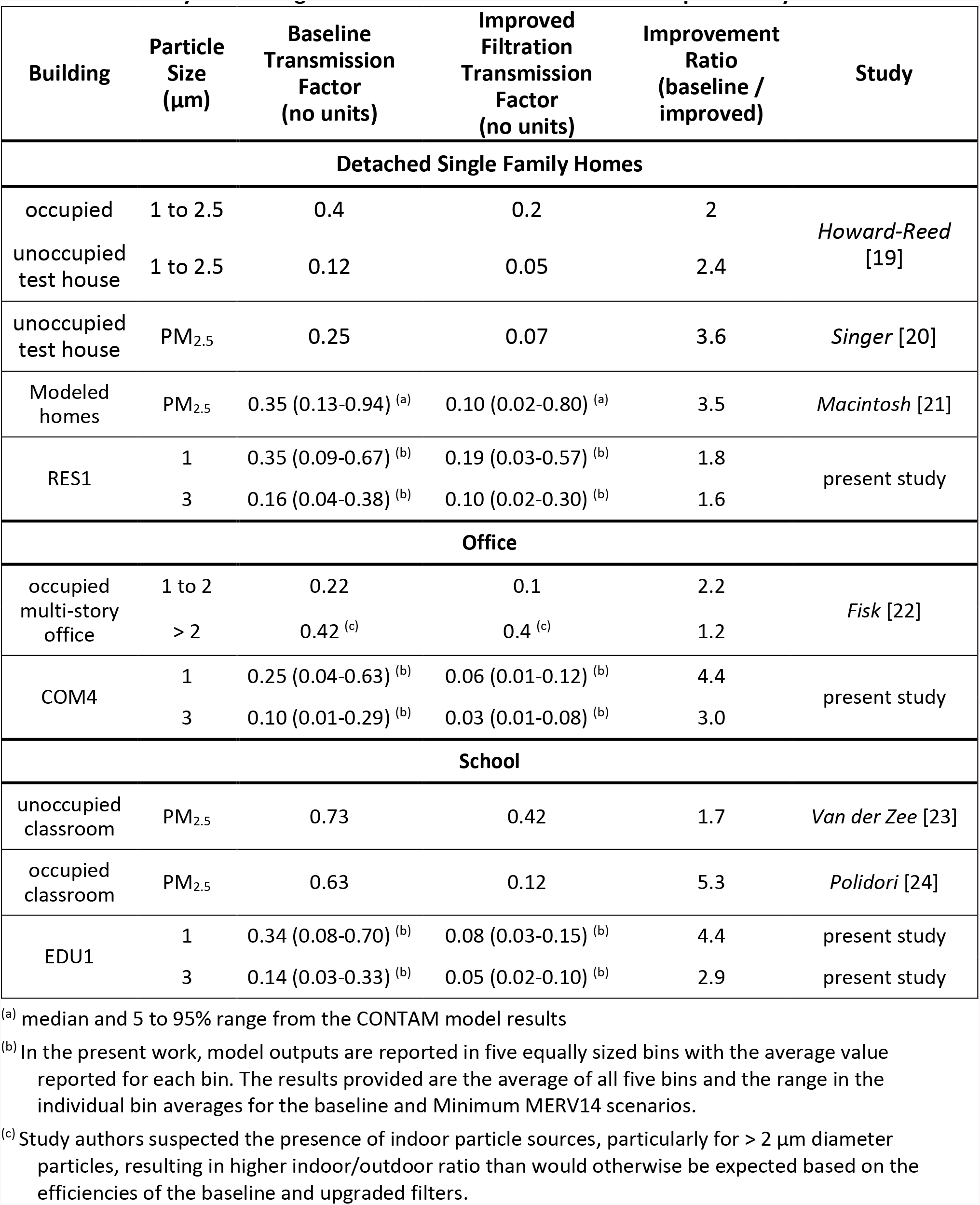
Summary of Building Transmission Factors for current and prior analyses.

*Singer et al*. [20] conducted an extensive set of ventilation and filtration system experiments in an unoccupied single family home. The reference furnace fan system contained a MERV 4 filter placed in the return duct to the air handler. The comparison system closest to the one modeled in the present work upgraded the MERV 4 filter to a higher particle removal efficiency (MERV 13) filter (system E in their work). The ratios of indoor to outdoor PM_2.5_ particle concentrations – based on the 24-hour average of several days of continuous data – are shown in **Table 11**. The observed indoor/outdoor PM_2.5_ concentration ratios are within the range of single family home Building Transmission Fractions predicted by the present study, which includes some systems with no filters. The observed improvement factor is larger than our modeling estimates.

*Macintosh et al*. [21] modeled the effectiveness of forced-air furnace-system air cleaning on reducing indoor concentrations of outdoor PM_2.5_. The analysis was based on the use of actual 24-hour average outdoor PM_2.5_ measurements during 2005 for three metropolitan areas in the state of Ohio, a set of single family prototypical dwelling designs, the CONTAM multizone air flow model, and two filtration assumptions. The first filtration assumption was a conventional furnace-system filter with an assumed PM_2.5_ removal efficiency of ∼14% (roughly equivalent to the MERV 5 filters used in the present analysis) and the second was an electrostatic precipitator (ESP) with an assumed filtration efficiency of ∼90% (an efficiency similar to the MERV 11–12 filtration category used here). The two systems were assumed to have different operating duty cycles – in the first, the forced air system operated only in response to heating/cooling demand while for the second, the system was assumed to have a variable speed fan that operated at high speed during heating/cooling demand and at half-speed during all other times. The simulation outputs, when combined across all prototypical buildings and the outdoor concentrations data from the three cities, yield the indoor/outdoor ratios shown in **Table 11**. The corresponding improvement factor is again similar to, but greater than, our modeling results.

Comparable tests in non-residential buildings are fewer. *Fisk et al*. [22] measured the ratio of indoor to outdoor particle concentrations in a large office building under two filtration scenarios: (a) the existing filter that had an estimated MERV rating of 5 to 6 and (b) a high efficiency filtration system corresponding to a MERV 15 filtration efficiency. The observed indoor/outdoor ratios for the two particle size categories are shown in **Table 11** along with our performance predictions for office buildings. The overall filtration efficiency and the improvement ratio observed by *Fisk et al*. are more modest than expected and are lower than our predictions. As discussed in [22], the experiments were conducted on two floors of the multistory building while both floors were occupied (160 to 280 people per floor). It is clear, especially from the data for particles larger than 2 µm in diameter, that there were important indoor sources for these particles that complicated the evaluation of the filtration performance (and improvement). As this building was selected to have non-smoking occupants, a likely, though unverified, source of larger particles was resuspension.

Improvements to particle filtration in schools have also been investigated. *Van der Zee et al*. [23] evaluated improving classroom filtration to a MERV 14 rated filter (from a baseline MERV 10 rated filter of dubious maintenance history) – along with other ventilation system performance changes. These changes resulted in a modest improvement in the indoor/outdoor PM_2.5_ ratio based on the non-teaching hour data (indoor particle concentrations during the teaching hours are strongly influenced by indoor particle sources), see **Table 11**. *Polidori et al*.[24] measured the airborne particle removal effectiveness of MERV 16 filters installed in HVAC systems serving classrooms in three schools – replacing baseline MERV 7 filters in these schools.^3^ The *Polidori et al*. study data collection was done entirely during school hours, though there were time periods during the day, such as outdoor recess and lunchtime, when the students were not in the classrooms. However, data during the empty classroom periods were not explicitly analyzed separately. The *Polidori et al*. measured ratio of real-time, concurrent indoor to outdoor PM_2.5_ concentrations demonstrated notably larger improvement with the increased filtration efficiency than the prior *Van der Zee et al*. study, see **Table 11**.

Our modeling predictions are broadly consistent with these school studies. Most of the observed Building Transmission Factor values are consistent with the upper bound of our modeling results. Also if the *van der Zee et al*. baseline filtration is MERV 10 (as appears to be the case) or even higher (if the existing, baseline filters were heavily loaded), then the observed improvement factor of 1.7× is similar to our calculated improvement factor of 2.5x and 1.5x for 1 and 3 µm diameter particles, respectively, using the Minimum MERV 11 scenario as the baseline filter. As seen in the analysis of single family homes, the improvement factor measured by *Polidori et al*. is similar to, but greater than, our modeling results.

### 6.2. Other Considerations

While our US building type filtration modeling results are broadly consistent with those of previous, individual building studies, the details of a particular building and/or the heating and cooling system will strongly influence the overall improvement seen for any given building. A recent analysis of residential filter effectiveness found that the “context” in which the filtration occurs matters. Context in this sense refers to a variety of factors that affect indoor air quality, including the overall building indoor/outdoor air change rate. Essentially this approach argues that filtration effectiveness is a system parameter that depends upon the overall operating conditions of the house [25]. For example, in some systems/houses they found greater filtration effectiveness in increasing system run time (furnace fan duty cycle) rather than simply upgrading the filter efficiency. This finding is consistent with the order of magnitude variability predicted by our study for residences where we explicitly account for variability in air change rates, indoor deposition losses, and, in the baseline scenario, furnace fan duty cycles. We note that the improved filtration scenarios examined in our analysis assume a 100% system run time (i.e., the furnace fan is always on).

The California Air Resources Board and US Environmental Protection Agency provide additional guidance on practical measures to improve residential air filtration, including the use of portable air filtration devices in buildings without forced air systems [26]–[28]. While a complete analysis of portable air filtration devices is beyond the scope of this study, we briefly discuss the use of portable air cleaners with regards to typical US residences.^4^ For discussion purposes, we use the Clean Air Delivery Rate (CADR) metric, which is the production rate of clean air (ft^3^ min^−1^) by a filtration system [30], [31].^5^ For typical residences with a MERV 7 rated filter and an always-on (100% duty cycle) furnace fan, the single family home and apartment furnace “CADR” values are ∼1200 and ∼440 ft^3^ min^−1^, respectively.^6^ In comparison, portable air cleaners intended for room cleaning have individual unit CADR ratings in the range of ∼250 to 400 ft^3^ min^−1^. Thus to achieve a comparable reduction in indoor airborne particle exposure, the use of one or more portable air cleaners would be needed. We note that individual portable air cleaners are typically intended to clean air in only one or two rooms, in contrast most furnace-based systems that are designed to distribute filtered air within the living space. Portable air cleaners can be useful alternatives for dwellings where forced air systems aren’t present, such as single family homes that lack forced air furnace systems or as a supplement to existing whole-house filtration, e.g., [32]. For context, the Association of Home Appliance Manufacturers (AHAM) recommends that “appropriately sized” portable air cleaners are 2/3 of the floor area of the space being cleaned [33], which corresponds to an additional loss term of 5 h^−1^ (λ_portable airfilter_ = CADR / Building Volume).

Another important simplifying assumption in our analysis is the use of a single, albeit well-mixed, box with closed windows and doors as the basis for modeling indoor particulate exposures. In the context of complex, multizone buildings; many larger buildings contain multiple HVAC systems which may vary in their filtration capabilities. In some cases, there is little air (and thus airborne particles) exchanged between zones and so the overall building can be modeled as the combination of independent, well-mixed zones as is done in this study with institutional housing (RES4, RES5, and RES6 building types). However, in other cases the effects of improved filtration may be physically localized in ways not captured in our estimates. As one example, we assume that HVAC filters in hospital air handling systems are uniformly rated MERV 14 or 15 and so, for this building type, no improvement is predicted (i.e., all hospitals already have the highest rated filters considered in our analysis). Future work could investigate the degree to which all zones have the same filtration capabilities and therefore if the air quality in some building zones could benefit from the use of higher rated filters.

Furthermore, the Indoor Normalized TSIAC results provided here are intended for use within a single-well mixed zone. As such they are not appropriate to assess within building, but cross zone contamination spread. For example, an apartment building may have many apartments, each with a separate HVAC system (zone). The Indoor Normalized TSIAC results presented here could be used to assess the degree to which particles released in one apartment exposes individuals in the same apartment. However, our Indoor Normalized TSIAC results, as calculated here, does not inform the degree to which individuals in a neighboring apartment are exposed.

Finally the parameters used in this study are representative descriptions of the factors affecting air flow and particle behavior in US buildings. In some cases, the parameter values chosen were based on limited information available in the literature and thus are thus based on the best existing data combined with our best engineering judgement. *Dillon et al*. [7] provides further discussion and suggestions for future research to improve these parameter values. We note that regardless of these uncertainties, our results broadly suggest that improving air filtration in buildings can reduce indoor airborne particle exposures across a wide range of building types. Further, the analysis methodology developed here could be applied to more detailed building models, including studies of specific buildings, where specific parameter values could be better characterized.

## 7. Acknowledgements

The authors also express their gratitude to their family for their support and enduring patience. The authors also thank Charles Dillon for his assistance during the development of this manuscript. Furthermore, the authors acknowledge the helpful reviews by Tarabay Antoun, Ron Baskett, Brooke Buddemeier, John Nasstrom, and Brenda Pobanz of the Lawrence Livermore National Laboratory; Dev Jani of the US Department of Homeland Security, Countering Weapons of Mass Destruction Office; Lance Wallace of Wallace Research; Woody Delp of the Lawrence Berkeley National Laboratory; and Thomas Phillips of Healthy Building Research.

This document was prepared as an account of work sponsored by an agency of the United States government. Neither the United States government nor Lawrence Livermore National Security, LLC, nor any of their employees makes any warranty, expressed or implied, or assumes any legal liability or responsibility for the accuracy, completeness, or usefulness of any information, apparatus, product, or process disclosed, or represents that its use would not infringe privately owned rights. Reference herein to any specific commercial product, process, or service by trade name, trademark, manufacturer, or otherwise does not necessarily constitute or imply its endorsement, recommendation, or favoring by the United States government or Lawrence Livermore National Security, LLC. The views and opinions of authors expressed herein do not necessarily state or reflect those of the United States government or Lawrence Livermore National Security, LLC, and shall not be used for advertising or product endorsement purposes.

Lawrence Livermore National Laboratory is operated by Lawrence Livermore National Security, LLC, for the U.S. Department of Energy, National Nuclear Security Administration under Contract DE-AC52-07NA27344.

## Data Availability

Supplemental material contains the data referred to in the manuscript

## 8. Attestations

### Ethics statement

No humans or animals were used this in work

### Data accessibility

No primary data were used in this analysis. The referenced supplemental material has been provided.

### Competing interests

The authors have no competing interests for the material provided in this manuscript beyond being and/or previously employed at our respective organizations.

### Author’s contributions

Michael Dillon (MBD) was responsible for the study concept and design, model development and analysis and manuscript drafting and revision.

Richard Sextro (RGS) provided quality control review of building metric equations, developed both the graphs and comparison against prior results, and significantly contributed to the overall manuscript drafting and revision.

Charles Dillon (CFD) provided editorial review and feedback during the manuscript revision process.

All authors give approval for release.

### Funding statement

MBD was supported, in part, by the US Department of Homeland Security for early related efforts.

RGS was self-supported.

CFD was self-supported.

## Auspices and Disclaimer

This work was performed under the auspices of the U.S. Department of Energy by Lawrence Livermore National Laboratory under Contract DE-AC52–07NA27344.

## Supplemental Material A: Detailed Results

The companion spreadsheet contains our modeling results for four filtration scenarios (baseline and three improvement scenarios) for each of the 32 building use types. The outputs for each filter and building combination include five aerosol sizes (0.1, 0.3, 1, 3, 10 µm diameter) and four airborne loss rates (0, 0.1, 1, 10 hr^−1^).

## Supplemental Material B: Results Summary

**Table B1.** Summary of US Building Transmission Factors – the ratio of the indoor exposures to the corresponding outdoor exposure – for selected building types, two aerosol sizes, and the 0 h^−1^ airborne loss rate. (dimensionless)

| Select Buildings<br>(Use Types) | Absolute Values |  |  |  | Relative Improvement from<br>Baseline Scenario |  |  |
| --- | --- | --- | --- | --- | --- | --- | --- |
|  | Baseline<br>Scenario | Min<br>MERV 7<br>Scenario | Min<br>MERV 11<br>Scenario | Min<br>MERV 14<br>Scenario | Min<br>MERV 7<br>Scenario | Min<br>MERV 11<br>Scenario | Min<br>MERV 14<br>Scenario |
| **1 µm aerodynamic diameter** |  |  |  |  |  |  |  |
| Single Family Homes (RES1) | 0.35 | 0.21 | 0.21 | 0.19 | 1.6 | 1.6 | 1.8 |
| Small Apt (RES3B, RES3C) | 0.18 | 0.055 | 0.054 | 0.034 | 3.2 | 3.2 | 5.1 |
| Large Apt (RES3E, RES3F) | 0.17 | 0.057 | 0.056 | 0.024 | 3.0 | 3.1 | 7.2 |
| Retail Stores (COM1) | 0.29 | 0.16 | 0.16 | 0.060 | 1.8 | 1.8 | 4.8 |
| Offices (COM4, COM5, GOV1) | 0.25 | 0.18 | 0.18 | 0.056 | 1.4 | 1.4 | 4.4 |
| Schools (EDU1) | 0.34 | 0.20 | 0.20 | 0.078 | 1.7 | 1.7 | 4.4 |
| **3 µm aerodynamic diameter** |  |  |  |  |  |  |  |
| Single Family Homes (RES1) | 0.16 | 0.10 | 0.10 | 0.10 | 1.6 | 1.6 | 1.6 |
| Small Apt (RES3B, RES3C) | 0.066 | 0.025 | 0.025 | 0.022 | 2.6 | 2.6 | 3.0 |
| Large Apt (RES3E, RES3F) | 0.052 | 0.020 | 0.020 | 0.015 | 2.5 | 2.6 | 3.4 |
| Retail Stores (COM1) | 0.11 | 0.055 | 0.055 | 0.036 | 2.0 | 2.0 | 3.1 |
| Offices (COM4, COM5, GOV1) | 0.10 | 0.059 | 0.057 | 0.032 | 1.6 | 1.7 | 3.0 |
| Schools (EDU1) | 0.14 | 0.073 | 0.071 | 0.048 | 1.9 | 2.0 | 2.9 |

**Table B2.** Summary of US Indoor Normalized Time and Space Integrated Air Concentrations (Indoor Normalized TSIAC) – indoor exposures to indoor-origin airborne particles – for selected building types, two aerosol sizes, and the 0 h^−1^ airborne loss rate. (s m^−1^)

| Select Buildings<br>(Use Types) | Absolute Values |  |  |  | Relative Improvement from<br>Baseline Scenario |  |  |
| --- | --- | --- | --- | --- | --- | --- | --- |
|  | Baseline<br>Scenario | Min<br>MERV 7<br>Scenario | Min<br>MERV 11<br>Scenario | Min<br>MERV 14<br>Scenario | Min<br>MERV 7<br>Scenario | Min<br>MERV 11<br>Scenario | Min<br>MERV 14<br>Scenario |
| **1 µm aerodynamic diameter** |  |  |  |  |  |  |  |
| Single Family Homes (RES1) | 18 | 11 | 10 | 9.8 | 1.7 | 1.7 | 1.8 |
| Small Apt (RES3B, RES3C) | 15 | 4.5 | 4.5 | 3.0 | 3.3 | 3.4 | 5.0 |
| Large Apt (RES3E, RES3F) | 9.6 | 4.0 | 4.0 | 2.6 | 2.4 | 2.4 | 3.7 |
| Retail Stores (COM1) | 6.3 | 4.8 | 4.7 | 3.7 | 1.3 | 1.3 | 1.7 |
| Offices (COM4, COM5, GOV1) | 3.3 | 3.0 | 2.9 | 2.4 | 1.1 | 1.1 | 1.4 |
| Schools (EDU1) | 5.0 | 4.1 | 4.0 | 3.2 | 1.2 | 1.2 | 1.5 |
| **3 µm aerodynamic diameter** |  |  |  |  |  |  |  |
| Single Family Homes (RES1) | 10 | 6.7 | 6.5 | 6.4 | 1.5 | 1.6 | 1.6 |
| Small Apt (RES3B, RES3C) | 7.2 | 2.8 | 2.8 | 2.6 | 2.6 | 2.6 | 2.8 |
| Large Apt (RES3E, RES3F) | 4.4 | 2.6 | 2.5 | 2.3 | 1.7 | 1.7 | 1.9 |
| Retail Stores (COM1) | 3.9 | 3.4 | 3.4 | 3.2 | 1.2 | 1.2 | 1.2 |
| Offices (COM4, COM5, GOV1) | 2.4 | 2.3 | 2.2 | 2.1 | 1.1 | 1.1 | 1.1 |
| Schools (EDU1) | 3.4 | 3.0 | 3.0 | 2.8 | 1.1 | 1.1 | 1.2 |

**Table B3.**
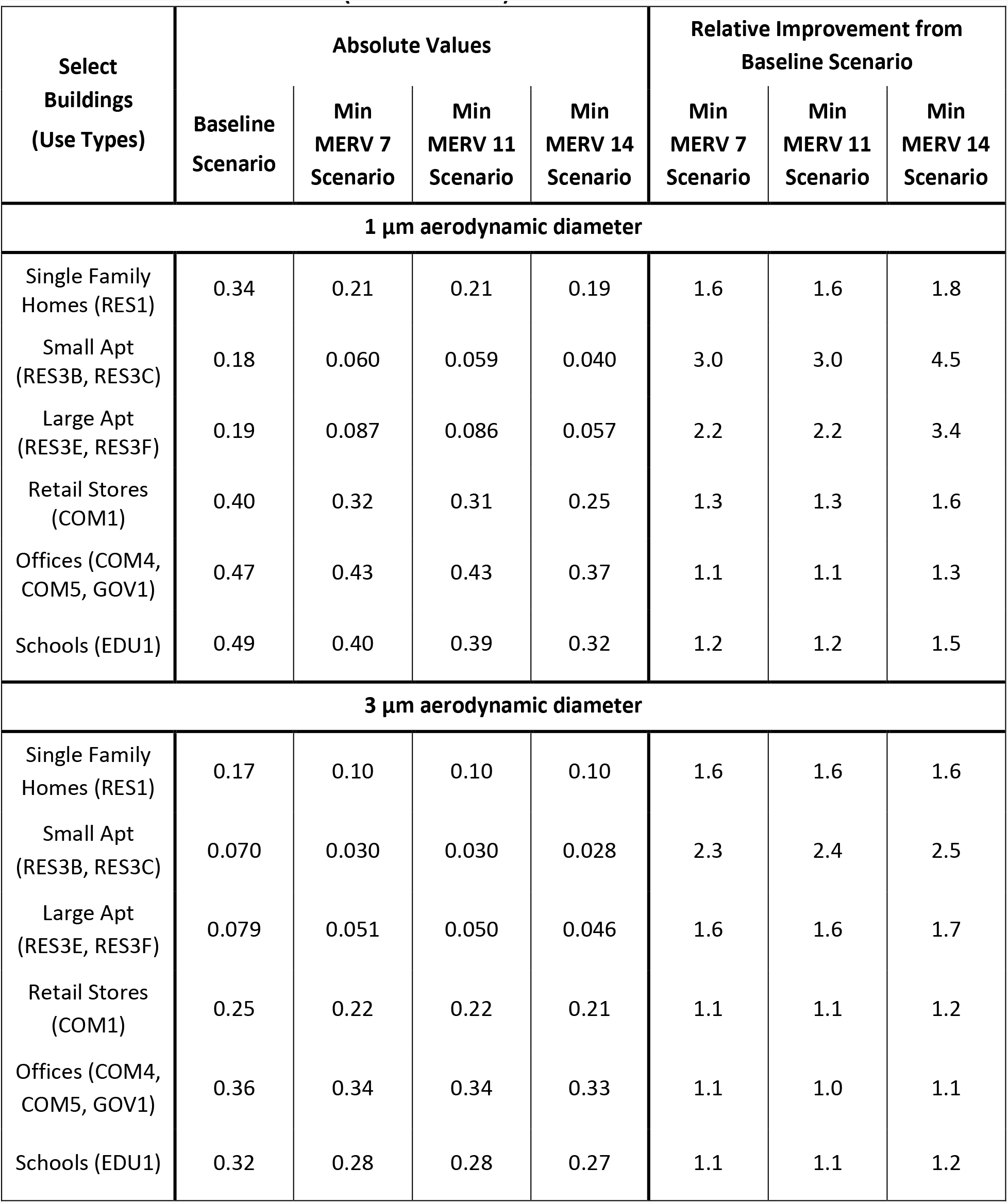
Summary of US Building Exit Fractions – the fraction of indoor airborne particles that are released to the outdoor atmosphere – for selected building types, two aerosol sizes, and the 0 h^−1^ airborne loss rate. (dimensionless)

| Select<br>Buildings<br>(Use Types) | Absolute Values |  |  |  | Relative Improvement from<br>Baseline Scenario |  |  |
| --- | --- | --- | --- | --- | --- | --- | --- |
|  | Baseline<br>Scenario | Min<br>MERV 7<br>Scenario | Min<br>MERV 11<br>Scenario | Min<br>MERV 14<br>Scenario | Min<br>MERV 7<br>Scenario | Min<br>MERV 11<br>Scenario | Min<br>MERV 14<br>Scenario |
| **1 µm aerodynamic diameter** |  |  |  |  |  |  |  |
| Single Family<br>Homes (RES1) | 0.34 | 0.21 | 0.21 | 0.19 | 1.6 | 1.6 | 1.8 |
| Small Apt<br>(RES3B, RES3C) | 0.18 | 0.060 | 0.059 | 0.040 | 3.0 | 3.0 | 4.5 |
| Large Apt<br>(RES3E, RES3F) | 0.19 | 0.087 | 0.086 | 0.057 | 2.2 | 2.2 | 3.4 |
| Retail Stores<br>(COM1) | 0.40 | 0.32 | 0.31 | 0.25 | 1.3 | 1.3 | 1.6 |
| Offices (COM4,<br>COM5, GOV1) | 0.47 | 0.43 | 0.43 | 0.37 | 1.1 | 1.1 | 1.3 |
| Schools (EDU1) | 0.49 | 0.40 | 0.39 | 0.32 | 1.2 | 1.2 | 1.5 |
| **3 µm aerodynamic diameter** |  |  |  |  |  |  |  |
| Single Family<br>Homes (RES1) | 0.17 | 0.10 | 0.10 | 0.10 | 1.6 | 1.6 | 1.6 |
| Small Apt<br>(RES3B, RES3C) | 0.070 | 0.030 | 0.030 | 0.028 | 2.3 | 2.4 | 2.5 |
| Large Apt<br>(RES3E, RES3F) | 0.079 | 0.051 | 0.050 | 0.046 | 1.6 | 1.6 | 1.7 |
| Retail Stores<br>(COM1) | 0.25 | 0.22 | 0.22 | 0.21 | 1.1 | 1.1 | 1.2 |
| Offices (COM4,<br>COM5, GOV1) | 0.36 | 0.34 | 0.34 | 0.33 | 1.1 | 1.0 | 1.1 |
| Schools (EDU1) | 0.32 | 0.28 | 0.28 | 0.27 | 1.1 | 1.1 | 1.2 |

**Table B4.** Summary of US Indoor Downwind Exposure Improvements for selected building types, two aerosol sizes, and the 0 h^−1^ airborne loss rate. (dimensionless)

| Select Buildings<br>(Use Types) | Relative Improvement from Baseline Scenario |  |  |
| --- | --- | --- | --- |
|  | Min MERV 7<br>Scenario | Min MERV 11<br>Scenario | Min MERV 14<br>Scenario |
| **1 µm aerodynamic diameter** |  |  |  |
| Single Family Homes (RES1) | 2.6 | 2.6 | 3.2 |
| Small Apt (RES3B, RES3C) | 9.6 | 9.8 | 23 |
| Large Apt (RES3E, RES3F) | 6.6 | 6.8 | 24 |
| **3 µm aerodynamic diameter** |  |  |  |
| Single Family Homes (RES1) | 2.5 | 2.6 | 2.6 |
| Small Apt (RES3B, RES3C) | 6.2 | 6.2 | 7.5 |
| Large Apt (RES3E, RES3F) | 4.0 | 4.2 | 5.8 |

## Supplemental Material C: Additional Building Metric Figures

**Figure C1** shows the Building Transmission Factors for the improved filtration scenarios. The indoor/outdoor exposure ratios are all smaller for the improved filtration scenarios than for the baseline case for the six building types, **Figure 1**, by the factors illustrated in **Figure 2**.

**Figure C1.**
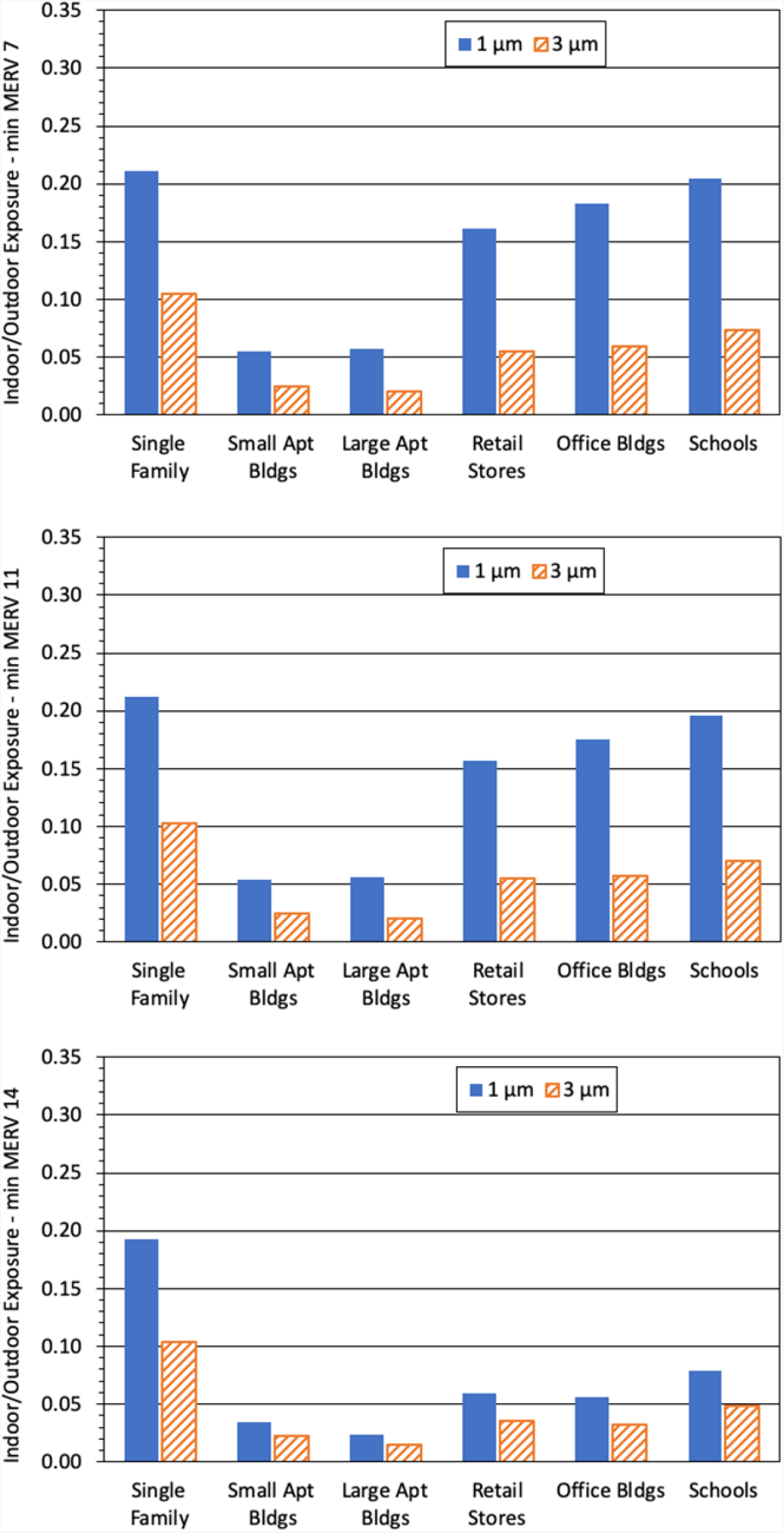
Improved filtration scenario results for the US Building Transmission Factor (dimensionless) – the ratio of the indoor exposures to the corresponding outdoor exposure – for selected building types; two aerosol sizes: 1 µm (solid bar) and 3 µm diameter (diagonal striped bar); and a 0 h^−1^ airborne loss rate. Higher Building Transmission Factor values correspond to greater exposures.

**Figure C2** shows the Indoor Normalized TSIAC for the improved filtration scenarios. The indoor exposure ratios are all smaller for the improved filtration scenarios than for the baseline case for the six building types, **Figure 3**, by the factors illustrated in **Figure 4**. Note the change in vertical scale from **Figure 3**.

**Figure C2.**
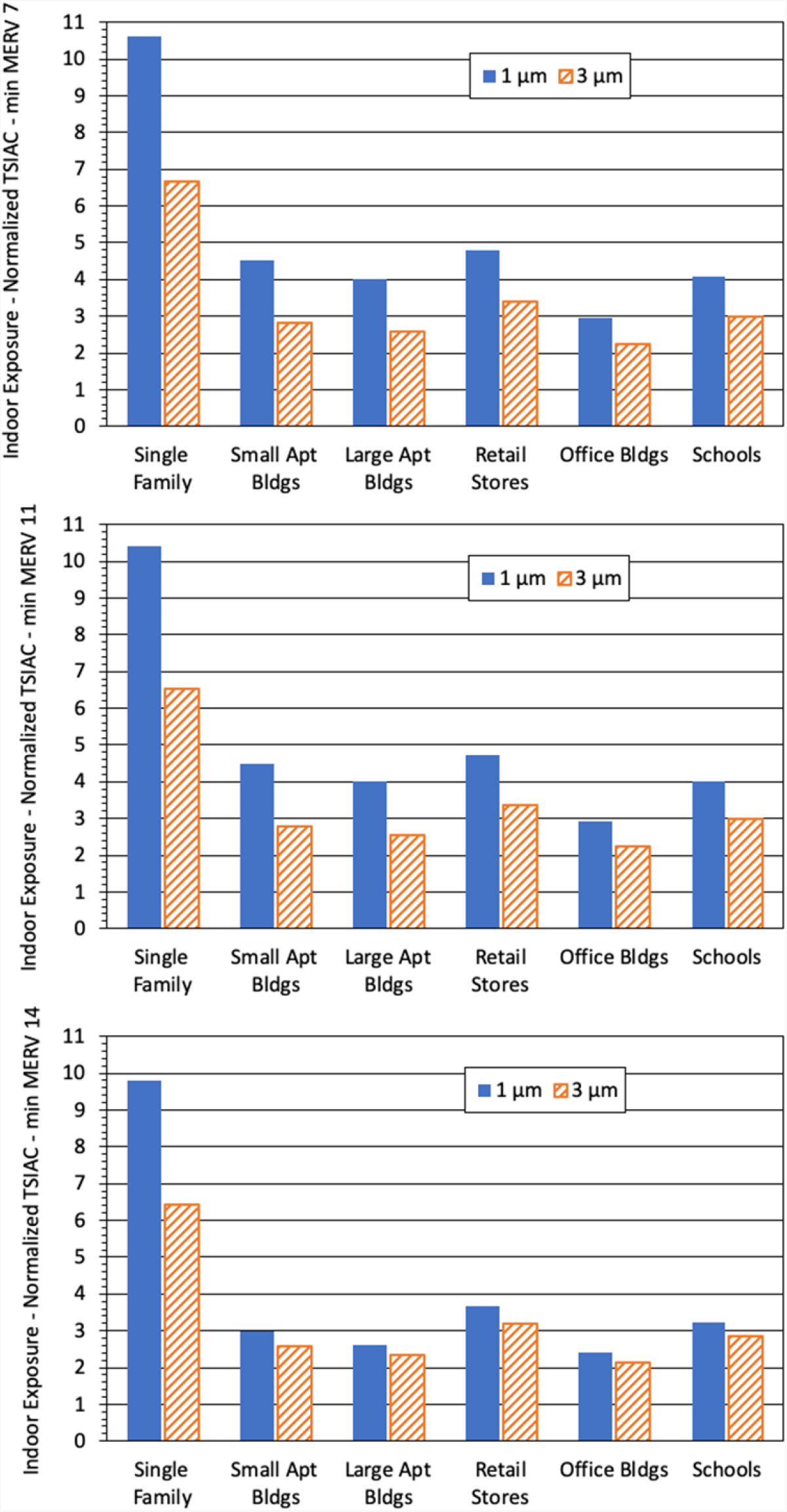
Improved filtration scenario results for the US Indoor Normalized Time and Space Integrated Air Concentration (Indoor Normalized TSIAC in s m^−1^) – indoor exposures to indoor-origin airborne particles – for selected building types; two aerosol sizes: 1 µm (solid bar) and 3 µm diameter (diagonal striped bar); and a 0 h^−1^ airborne loss rate. Higher Indoor Normalized TSIAC values correspond to greater exposures. Note the change in vertical scale from Figure 3.

**Figure C3** shows the Building Exit Fraction for the improved filtration scenarios. The Building Exit Fractions are all smaller for the improved filtration scenarios than for the baseline case for the six building types, **Figure 5**, by the factors illustrated in **Figure 6**.

**Figure C3.**
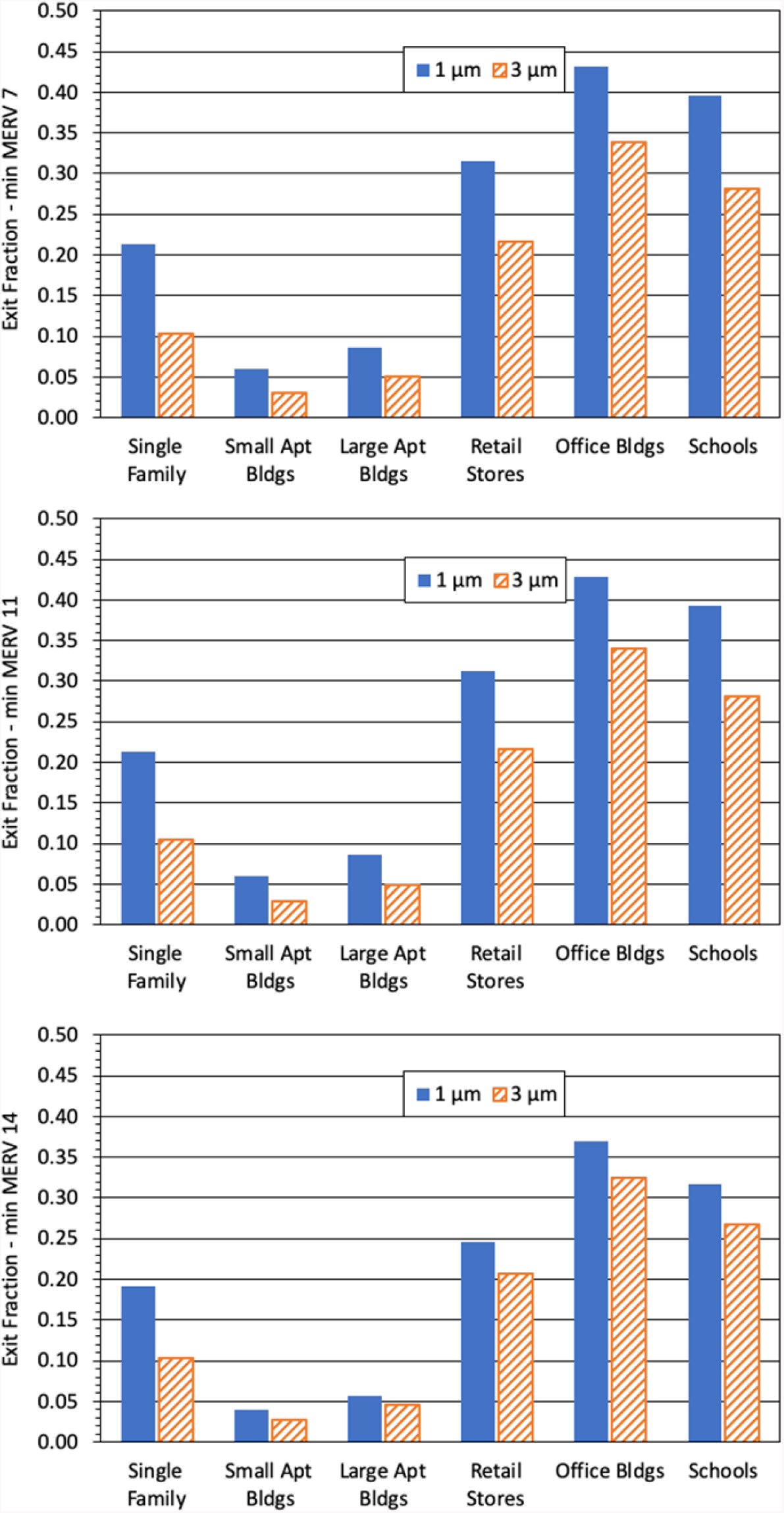
Improved filtration scenario results for the US Building Exit Fractions (dimensionless) – the fraction of indoor airborne particles that are released to the outdoor atmosphere – for selected building types; two aerosol sizes: 1 µm (solid bar) and 3 µm diameter (diagonal striped bar); and a 0 h^−1^ airborne loss rate. Higher Exit Fraction values correspond to more particles released to the outdoor atmosphere.

## Supplemental Material D: Improvement Sensitivity to Particle Size and Airborne Loss Rate

**Figures D1** to **D3** illustrates how the improvements to (reduction in) indoor airborne particle exposure varies with airborne loss rate and particle size using the Building Transmission Factor metric, the Minimum MERV 14 scenario, and selected building types. Broadly, the improvement decreases with increasing “generic” first-order airborne loss rate, λ_*decay*_ and particle size. The small and large apartment building improvements are sensitive to airborne loss rate and particle size. The single family home, retail store, office building and school improvements are notably less sensitive.

**Figure D1.**
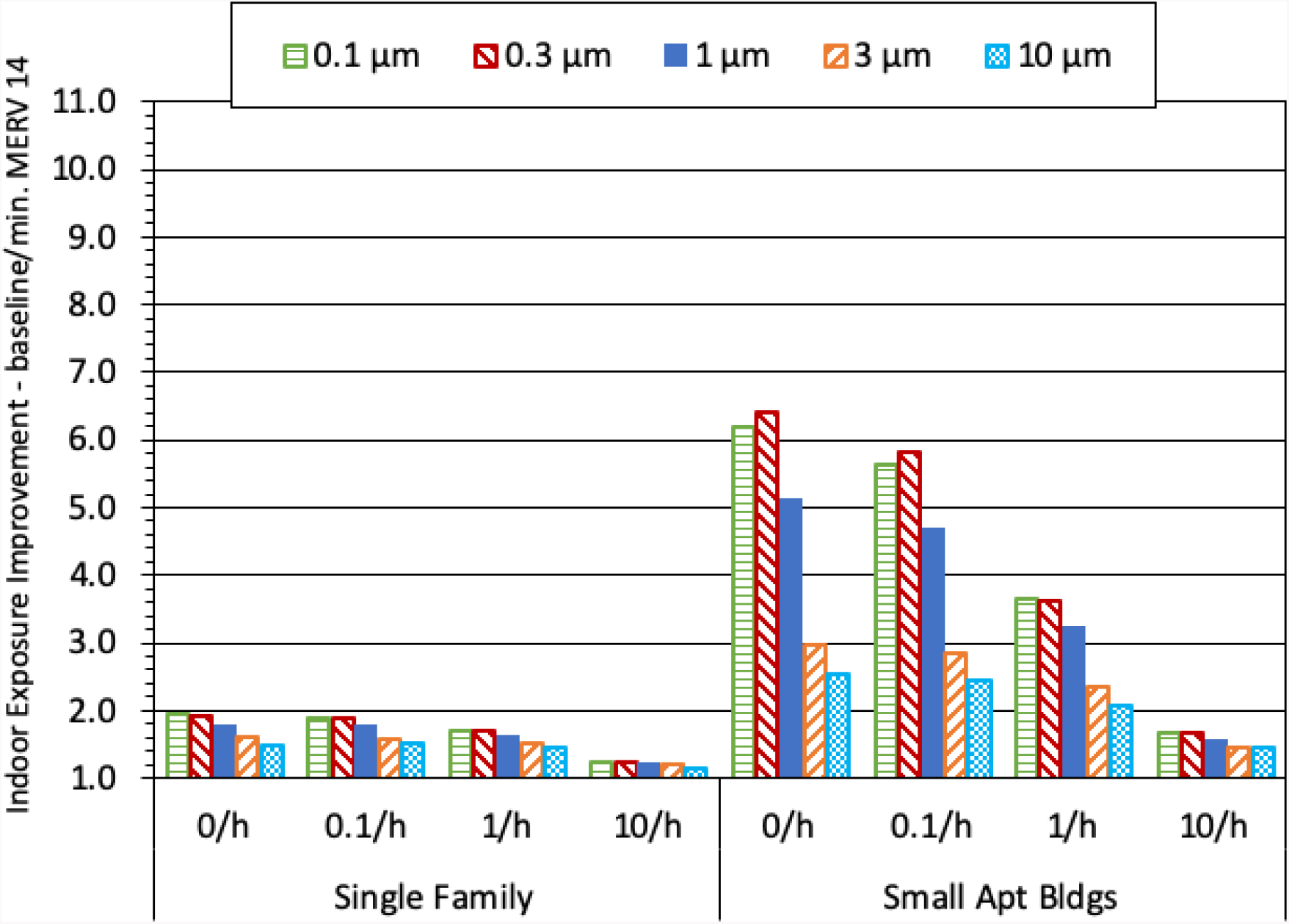
Improvement in the US Building Transmission Factor for two residential building types; five aerosol sizes: 0.1 µm (horizontal striped bar), 0.3 µm (downward diagonal striped bar, 1 µm (solid bar), 3 µm diameter (upward diagonal striped bar), and 10 µm diameter particles (cross-hatched bar); and four airborne loss rates: 0, 0.1, 1, and 10 h^−1^. The vertical axis is the ratio of the Building Transmission Factors for the baseline to the Minimum MERV 14 filtration scenario (a value of 1 indicates no improvement, larger numbers indicate greater improvement).

**Figure D2.**
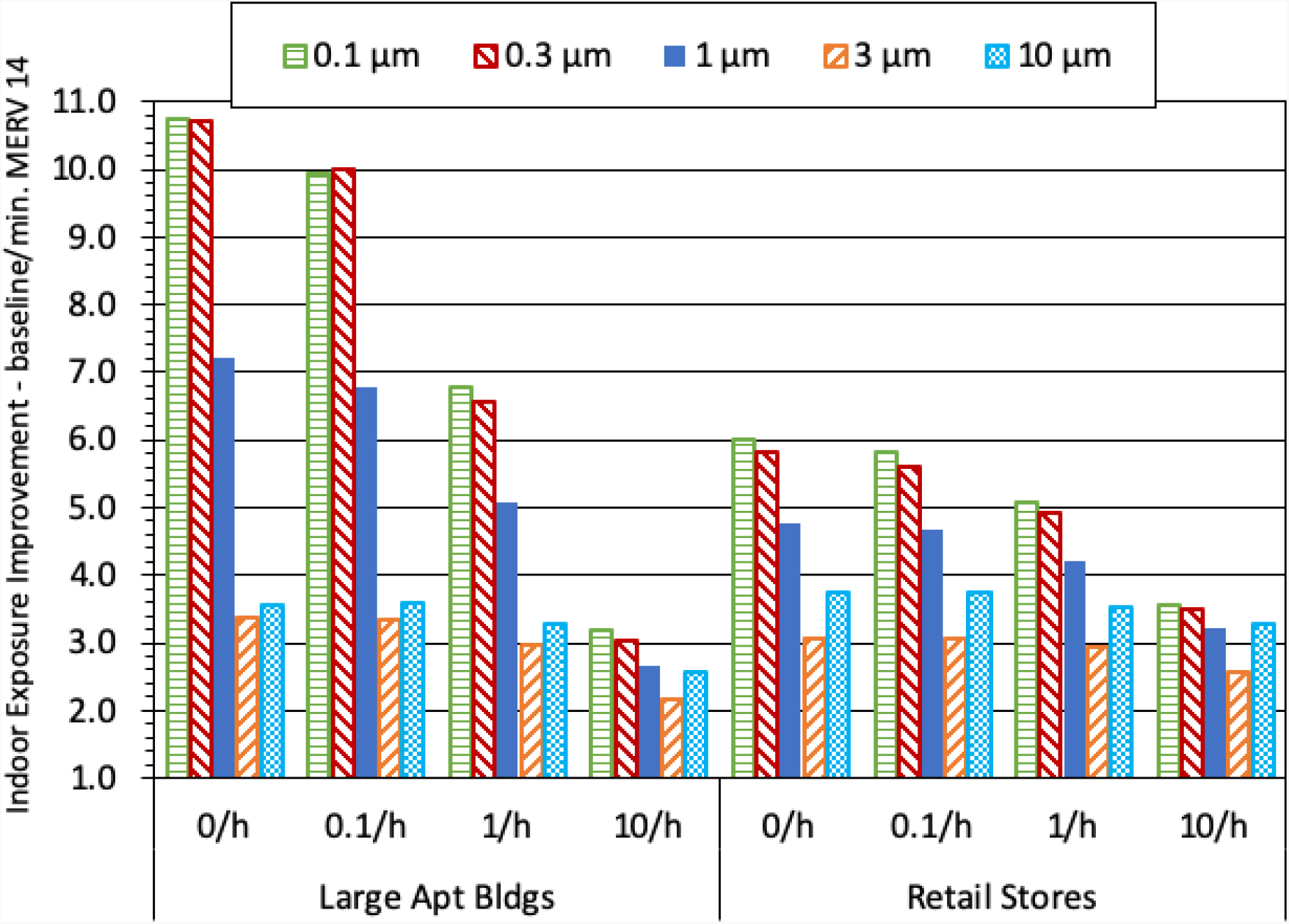
Improvement in the US Building Transmission Factor for large apartments and retail stores; five aerosol sizes: 0.1 µm (horizontal striped bar), 0.3 µm (downward diagonal striped bar, 1 µm (solid bar), 3 µm diameter (upward diagonal striped bar), and 10 µm diameter particles (cross-hatched bar); and four airborne loss rates: 0, 0.1, 1, and 10 h^−1^. The vertical axis is the ratio of the Building Transmission Factors for the baseline to the Minimum MERV 14 filtration scenario (a value of 1 indicates no improvement, larger numbers indicate greater improvement).

**Figure D3.**
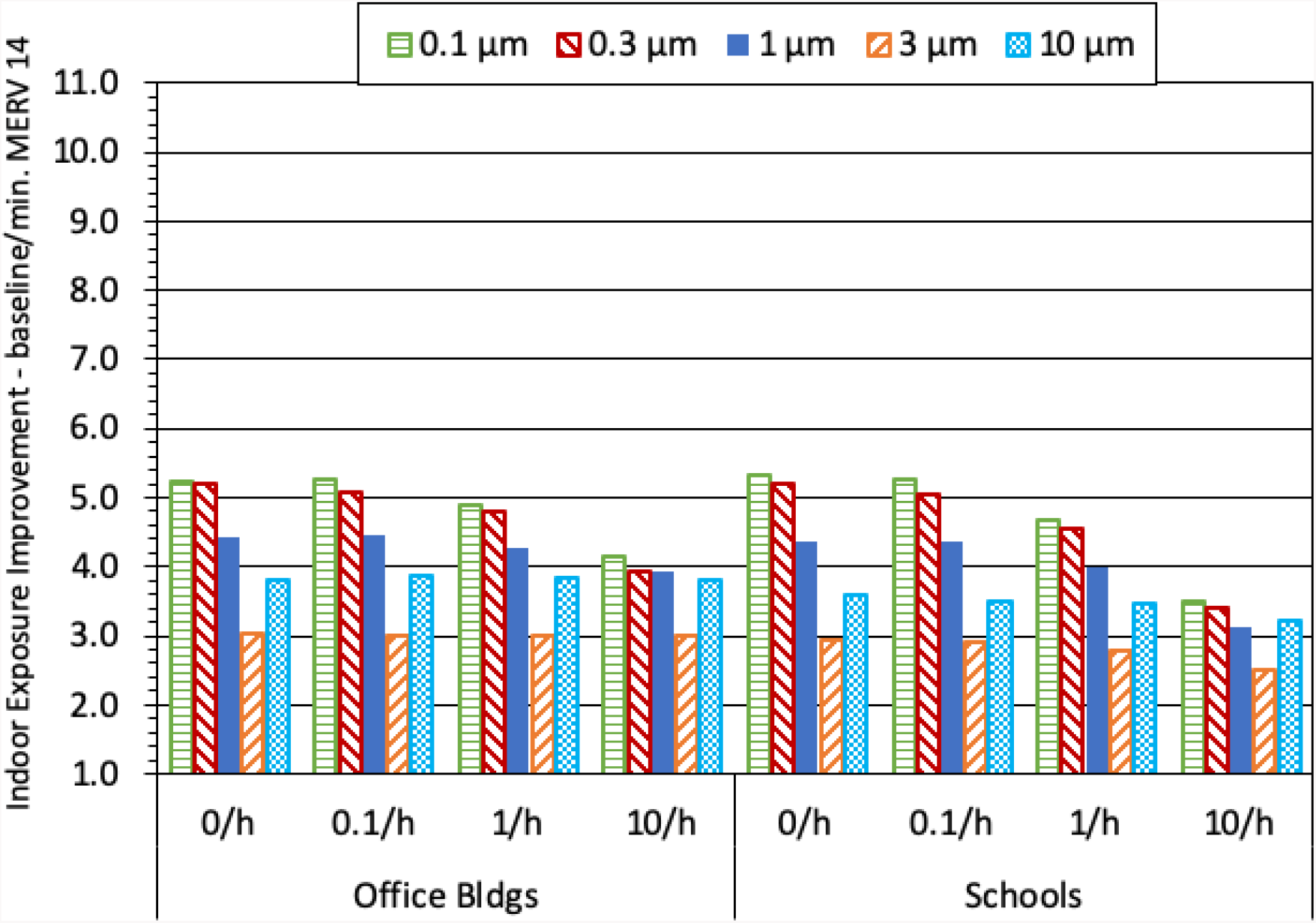
Improvement in the US Building Transmission Factor for office buildings and schools; five aerosol sizes: 0.1 µm (horizontal striped bar); 0.3 µm (downward diagonal striped bar, 1 µm (solid bar), 3 µm diameter (upward diagonal striped bar), and 10 µm diameter particles (cross-hatched bar); and four airborne loss rates: 0, 0.1, 1, and 10 h^−1^. The vertical axis is the ratio of the Building Transmission Factors for the baseline to the Minimum MERV 14 scenario (a value of 1 indicates no improvement, larger numbers indicate greater improvement).

## Footnotes

1 The mean of the “Median,” “2^nd^ Lowest,” and “Lowest” distribution categories is 0.06 for the Minimum MERV 14 scenario; RES1 building use type; 1 um diameter particles; and 0 hr^−1^ airborne loss rate. For context, the mean of all distribution categories is 0.19. The corresponding mean for the baseline scenario is 0.35. See **Supplemental Material A: Detailed Results** for more details.

2 Data from this study was used in developing the indoor deposition rate values used in our analysis.

3 Data from this study was one of the data sources used in developing the airflow parameter values used in our analysis.

4 Per [29], the typical US detached single family home and apartment has 2,200 and 840 ft^2^ of heated floor area, respectively. Assuming an 8 ft room height, the corresponding building volumes are 18,000 and 6,700 ft^3^, respectively.

5 We note that the formal AHAM CADR *testing standard* does not cover the whole-building filtration systems but the term can be applied to any forced air filtration system.

6 CADR = *F_filter_* x *F_r,fan_* × *r_fan_* × *Building Volume*. Per **Table 3**, the furnace fan flow rate is 5.7 h^−1^ and per **Table 10**, the median MERV 7 rated filter efficiency for 1 µm particles is 0.69.

## Notes

### Competing Interest Statement

The authors have declared no competing interest.

### Funding Statement

MBD was supported, in part, by the US Department of Homeland Security. Outside the scope of this work, MBD collaborates with the University of Texas Medical Branch.

